# Adjuvanted RSVPreF3 vaccine impact over 3 RSV seasons in older adults with comorbidities

**DOI:** 10.64898/2026.04.09.26348324

**Authors:** Alberto Papi, David M. G. Halpin, Robert G. Feldman, Michael G. Ison, Tino F. Schwarz, Dong-Gun Lee, Raffaele Antonelli Incalzi, Laurence Fissette, Stebin Xavier, Marie-Pierre David, Jean-Philippe Michaud, Shady Kotb, Céline Maréchal, Aurélie Olivier, Veronica Hulstrøm, Marie Van der Wielen, the AReSVi-006 study group

**Affiliations:** Pulmonary Division, University of Ferrara, St. Anna University Hospital, Ferrara, Italy; Department of Health and Community Sciences, University of Exeter Medical School, Exeter, United Kingdom; Senior Clinical Trials, Inc, Laguna Hills, CA, United States; Bethesda, MD, United States; Institute of Laboratory Medicine and Vaccination Center, Klinikum Würzburg Mitte, Campus Juliusspital, Würzburg, Germany; Division of Infectious Diseases, Department of Internal Medicine, Vaccine Bio Research Institute, College of Medicine, The Catholic University of Korea, Seoul, South Korea; Fondazione Policlinico Universitario Campus Bio-Medico e Università Campus Bio-Medico, Rome, Italy; GSK, Wavre, Belgium; GSK, Bengaluru, India

**Keywords:** RSV, vaccination, efficacy, chronic obstructive pulmonary disease, asthma, diabetes, obesity

## Abstract

**Background:** We explored the efficacy of AS01_E_-adjuvanted respiratory syncytial virus prefusion F protein-based vaccine (adjuvanted RSVPreF3) in subpopulations of participants with underlying medical conditions in the multi-country, phase 3 AReSVi-006 trial (conducted May/2021-May/2024).

**Methods:** Medically stable ≥60-year-olds were 1:1-randomised to receive one adjuvanted RSVPreF3 or placebo dose pre-RSV season 1. In exploratory post-hoc analyses in subgroups of participants with underlying conditions (including COPD, asthma, diabetes, obesity [BMI≥30 kg/m^2^]), we evaluated efficacy of one vaccine dose against RSV-related lower respiratory tract disease (RSV-LRTD), acute respiratory illness (RSV-ARI), and RSV-ARI-related complications (e.g., pneumonia, COPD/asthma exacerbation, cardiovascular events). We also evaluated (post-hoc) RSV-ARI-related systemic corticosteroid and antibiotics use in participants with COPD or asthma.

**Results:** The efficacy analyses comprised 12,468 vaccine and 12,498 placebo recipients. Efficacy against RSV-LRTD over three RSV seasons was similar among participants with COPD (75.1%, 95% CI: 40.2-91.4), asthma (65.8%, 31.0-84.7), diabetes (69.8%, 37.5-87.1), and obesity (74.1%, 56.4-85.5) as in the overall study population (62.9%, 97.5% CI: 46.7-74.8). Efficacy was also observed against RSV-ARI in these subgroups. Efficacy against RSV-ARI-related complications was 74.4% (95% CI: 11.2-95.2) in participants with COPD and 60.8% (−9.9-88.7) in those with asthma. Among participants with COPD, 15.4% (1.9-45.4) of RSV-ARI episodes in vaccine vs 22.4% (12.5-35.3) in placebo recipients were treated with systemic corticosteroids, and 46.2% (19.2-74.9) vs 56.9% (43.2-69.8) with antibiotics.

**Conclusions:** Post-hoc analyses of the AReSVi-006 trial suggest that adjuvanted RSVPreF3 may help prevent RSV-ARI, RSV-LRTD, and RSV-related complications in medically stable older adults with underlying medical conditions like COPD and asthma.

**Trial registration:** ClinicalTrials.gov: NCT04886596

**Summary:** Post-hoc analyses of the AReSVi-006 trial suggest that 1 dose of adjuvanted RSVPreF3 may help prevent RSV-related illness and complications over 3 consecutive RSV seasons in subgroups of ≥60-year-olds with chronic medical conditions, e.g., COPD and asthma.

## Background

Respiratory syncytial virus (RSV) is an important cause of acute respiratory illness (ARI). Adults aged ≥60 years and those with underlying medical conditions, such as asthma, chronic obstructive pulmonary disease (COPD), congestive heart failure, diabetes, severe obesity, and kidney and liver disorders have an elevated risk of severe outcomes from RSV, including hospitalisation and death [1–7]. In these populations, RSV infection can lead to exacerbations of underlying COPD or asthma and other complications, such as pneumonia and acute cardiac events [6–12]. Exacerbations of obstructive respiratory diseases are characterised by acute worsening of respiratory symptoms that may result in an increased risk of future severe exacerbations, accelerated loss of lung function, disease progression, and death [13–15].

Several vaccines for the prevention of RSV-related lower respiratory tract disease (RSV-LRTD) have recently become available [16]. The AS01_E_-adjuvanted RSV prefusion F protein-based vaccine (adjuvanted RSVPreF3, *Arexvy*, GSK) has been licensed in many countries for adults aged ≥60 years and for those aged 50−59 years at increased risk of (severe) RSV disease [17,18]. Vaccine efficacy against RSV-LRTD and RSV-ARI in participants aged ≥60 years was shown in the AReSVi-006 phase 3 trial [19–21]. The vaccine was also efficacious among participants with ≥1 chronic medical condition known to increase the risk of severe RSV disease [19–22]. Moreover, real-world case-control studies have shown that RSV vaccines, including adjuvanted RSVPreF3, are effective against RSV-related ARI, hospitalisations, and urgent care visits in ≥60-year-olds [23–26].

Considering the high RSV burden in individuals with underlying medical conditions, we further explored the efficacy of adjuvanted RSVPreF3 in this population. We focused on conditions that increase the risk for severe RSV disease that were most prevalent in the AReSVi-006 trial population (COPD, asthma, diabetes, and obesity) and evaluated efficacy against RSV-LRTD and RSV-ARI in these subgroups and efficacy against RSV-ARI-related complications (including COPD and asthma exacerbations). We also assessed the use of systemic corticosteroids and antibiotics to treat RSV-ARI in participants with COPD and asthma in vaccinated versus unvaccinated participants. Findings are summarised in plain language in **Figure 1**.

**Figure 1.**
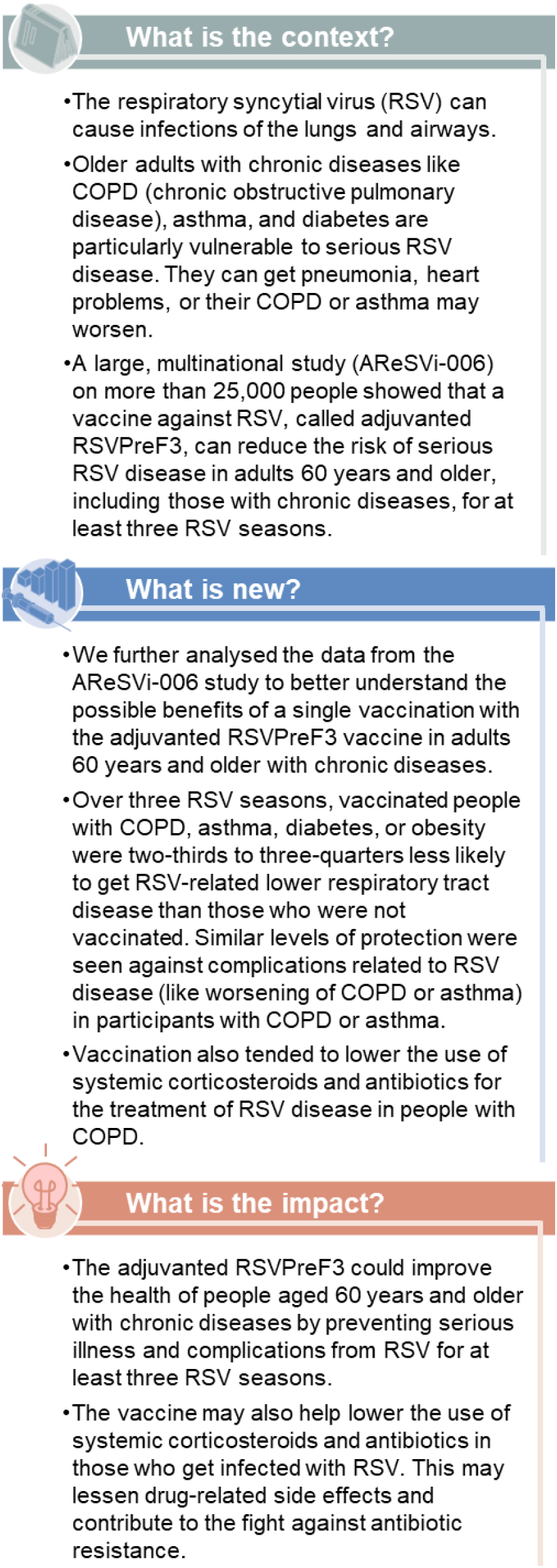
Plain language summary.

## Methods

### Study design, participants, and procedures

This randomised, placebo-controlled, observer-blind, phase 3 trial (ClinicalTrials.gov: NCT04886596) was conducted between May 2021 and May 2024 in 17 countries. The trial sites’ independent ethics committees approved the protocol (available on https://www.gsk-studyregister.com/en/trial-details/?id=212494) and other study-related documents. The trial was performed in accordance with the Declaration of Helsinki, Good Clinical Practice, and local laws and regulations. Participant safety was monitored by an independent data and safety monitoring committee.

Results for the primary confirmatory objective (efficacy of a single dose against RSV-LRTD during RSV season 1), secondary confirmatory objectives (efficacy of a single dose and of a first dose followed by a second dose 1 year later against RSV-LRTD over two and three RSV seasons), and secondary descriptive objectives were published before [19–22,27]. Here, we report results from exploratory (mostly post-hoc) analyses in participants with pre-existing medical conditions.

Detailed methods were described previously [19–22]. Medically stable ≥60-year-olds, including individuals with chronic stable medical conditions with or without specific treatment, were enrolled after providing written or witnessed informed consent. Participants were randomised (1:1) before RSV season 1 to receive a single intramuscular dose of vaccine (adjuvanted RSVPreF3 group) or placebo (placebo group). Before RSV season 2, participants who had received vaccine were re-randomised (1:1) to receive a second vaccine dose (RSV revaccination group) or placebo (RSV single-dose group). Individuals in the placebo group received another placebo dose pre-season 2 (**Figure 2**). The trial initially planned for another adjuvanted RSVPreF3 or placebo dose to be administered before season 3. However, because results after two seasons showed no benefit from a second dose compared to a single dose in the overall trial population [20], the protocol was amended to remove the pre-season 3 dose. A subset of participants received the pre-season 3 dose before the amendment was approved (**Figure 2**) [21].

**Figure 2.**
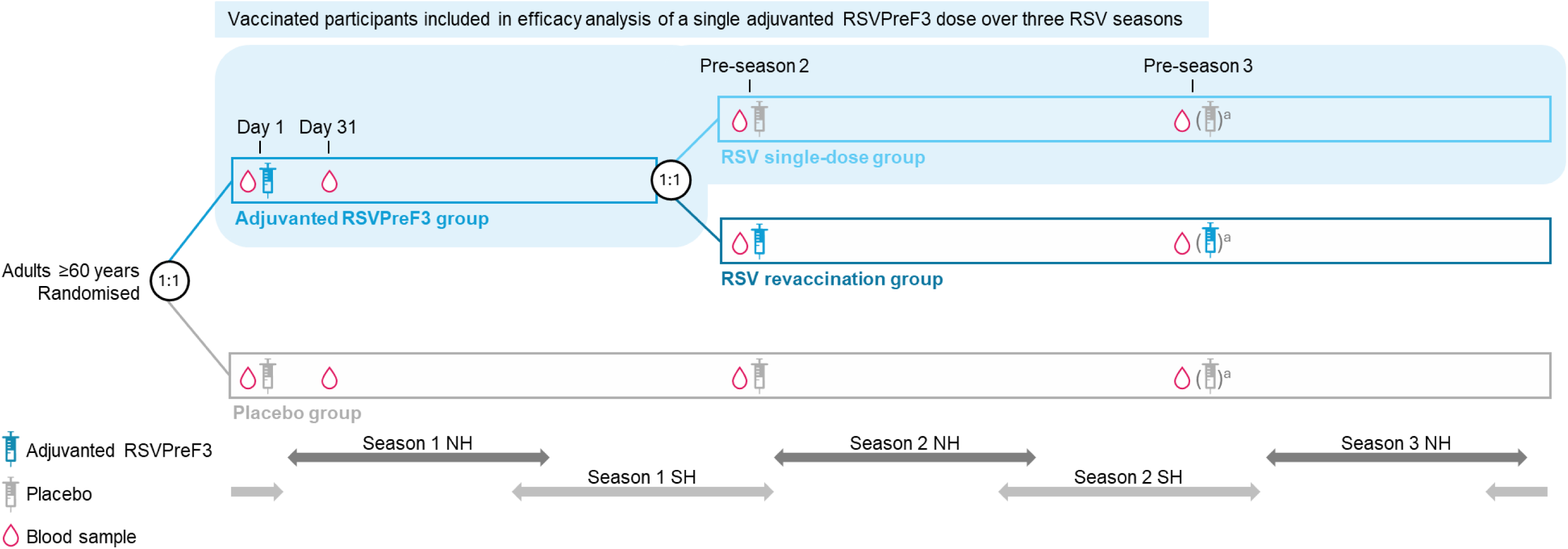
Study design. Participants were randomised to receive a dose of adjuvanted respiratory syncytial virus (RSV) prefusion F protein-based vaccine (adjuvanted RSVPreF3) or placebo pre-RSV season 1. Participants in the adjuvanted RSVPreF3 group were re-randomised to receive a second vaccine dose (RSV revaccination group) or placebo (RSV single-dose group) pre-season 2. Participants in the placebo group received a second placebo dose pre-season 2. RSV seasons were defined in the protocol as from 1 October to 30 April in the Northern Hemisphere (NH) and from 1 March to 30 September in the Southern Hemisphere (SH). The blue shaded area on the figure shows the vaccinated participants who were included in the efficacy analysis of a single vaccine dose over three seasons. ^a^After the trial’s season 2 analysis [20], the protocol was amended to remove the dose originally planned to be administered pre-season 3. For 5742 participants (RSV single-dose: 1467, RSV revaccination: 1409, placebo: 2866), the pre-season 3 visit took place before the amendment was approved; hence, they received adjuvanted RSVPreF3 or placebo pre-season 3 [21].

Information regarding pre-existing medical conditions was collected through participant interviews and medical record review at baseline. Medical conditions of interest used for subgroup analyses were those associated with an elevated risk of severe RSV disease and were divided into cardiorespiratory conditions (any chronic respiratory/pulmonary disease, including COPD and asthma, and chronic heart failure) and endocrine or metabolic conditions (type 1 or 2 diabetes and advanced liver or kidney disease). A pre-defined list of preferred terms from the Medical Dictionary for Regulatory Activities was used to identify the relevant medical conditions in the database. Participant characteristics, including body mass index (BMI), were assessed at baseline. Obesity was defined as a BMI ≥30 kg/m^2^.

Participants were followed up for the occurrence of ARIs during three RSV seasons in the Northern Hemisphere and at least two seasons in the Southern Hemisphere (**Figure 2**). ARI surveillance was performed via spontaneous reporting by the participants and scheduled site staff contacts [19]. Whenever a participant exhibited two or more ARI symptoms or signs, an assessment visit was required, during which staff examined the participant and collected nasal and throat swabs. Participants also performed nasal self-swabs [19]. Both swab types were used to detect RSV-A and RSV-B through quantitative reverse transcriptase-polymerase chain reaction. ARI was characterised by the presence of at least two respiratory symptoms or signs, or at least one respiratory and one systemic symptom or sign, persisting ≥24 hours. LRTD was defined as having at least two lower respiratory symptoms or signs (including at least one lower respiratory sign) or at least three lower respiratory symptoms, persisting ≥24 hours; LRTD was classified as severe if clinical signs and/or investigator evaluation indicated severe disease, or if supportive therapy was required (**Supplementary methods**) [19]. Only RSV-LRTD cases validated as LRTD by an adjudication committee were included in the analyses [19].

At each study visit/contact and during the ARI assessment visit, any medication (prescribed or self-medication) taken to treat an ARI or ARI-related complication was recorded in the electronic case report form with medical indication.

Complications were collected throughout the study and recorded in the electronic case report form. They included respiratory complications (pneumonia, new diagnosis or exacerbation of COPD or asthma, other respiratory complications [e.g., sinusitis, bronchitis, dyspnoea]) and non-respiratory complications (new-onset or worsening congestive heart failure, myocardial infarction, stroke, diabetes, other non-respiratory complications [e.g., otitis media]) (**Supplementary methods**). The relationship of these complications to an RSV-ARI episode was assessed by the investigators based on their clinical judgment.

Immunogenicity assessments are described in the **Supplementary methods**.

### Statistical analyses

The planned sample size was 25,000 participants [19–21]. Analyses were performed with SAS Data Science 3.6 (SAS Institute, Cary, NC, USA). All analyses described here were exploratory with no adjustments for multiplicity.

Efficacy analyses were performed in the modified exposed population (participants who received a vaccine or placebo dose and did not report RSV-ARI within 15 days post-dose 1). Efficacy against first occurrence of RSV-LRTD and RSV-ARI by pre-existing medical condition of interest were pre-specified secondary and tertiary endpoints, respectively, and were analysed in subgroups of participants without any of the pre-existing conditions of interest, with ≥1 condition, ≥1 cardiorespiratory, and ≥1 endocrine or metabolic condition of interest. Efficacy against first occurrence of RSV-ARI-related complications in the overall study population was also a pre-specified secondary endpoint. Subgroup analyses of the efficacy against RSV-LRTD, RSV-ARI, and RSV-ARI-related complications by individual pre-existing conditions of interest (e.g., COPD, asthma, diabetes) and by BMI category were performed post hoc, as were efficacy analyses against severe RSV-LRTD by pre-existing conditions of interest. In addition, the frequencies (calculated with exact 95% CIs) of RSV-ARI episodes treated with systemic corticosteroids or antibiotics were analysed post hoc, overall and by pre-existing condition of interest.

To evaluate the cumulative efficacy of adjuvanted RSVPreF3 over three RSV seasons, the time at risk started on day 15 post-dose 1 and ended at the first occurrence of an event, drop-out, end of the third Northern Hemisphere season (30 April 2024), or end of the first Southern Hemisphere season (30 September 2022) for participants who did not receive dose 2. Participants who received an RSV vaccine not planned per protocol were censored at the receipt of that vaccine. To evaluate the efficacy of a single vaccine dose, participants in the RSV single-dose group and in the placebo group contributed to the entire follow-up, whereas participants in the RSV revaccination group contributed to season 1 but were censored at dose 2 (**Figure 2**). Efficacy was defined as 1 − incidence rate ratio (vaccine vs placebo group), calculated with exact confidence intervals (CIs) using the conditional exact binomial method based on a Poisson model adjusted by age category, region, and season.

## Results

### Study population

A total of 24,972 participants were included in the exposed population, with 12,469 in the adjuvanted RSVPreF3 group and 12,503 in the placebo group. Of these, 24,966 participants (adjuvanted RSVPreF3: 12,468, placebo: 12,498) were part of the modified exposed population, and 18,726 (75.0%; adjuvanted RSVPreF3: 9365, placebo: 9361) completed the study. Reasons for withdrawal and important protocol deviations were reported previously [21].

Baseline characteristics were balanced between study groups [21]. In total, 5014 (40.2%) participants in the vaccine group and 4954 (39.6%) in the placebo group had ≥1 pre-defined pre-existing condition of interest (i.e., associated with an increased risk of RSV disease); 2577 (20.7%) and 2505 (20.0%), respectively, had ≥1 cardiorespiratory condition and 3243 (26.0%) and 3276 (26.2%), respectively, had ≥1 endocrine/metabolic condition. The mean age was 70.2 years in both groups among participants with ≥1 condition of interest and 69.1 years among those with none of the conditions. The mean BMI was 30.5 kg/m^2^ in both groups among participants with ≥1 condition of interest and 28.2 kg/m^2^ among those with none of the conditions (**Table 1**).

**Table 1.** Baseline characteristics of participants with or without pre-existing condition of interest (exposed population)

| Characteristic | ≥1 pre-existing condition of interest |  | No pre-existing conditions of interest |  |
| --- | --- | --- | --- | --- |
|  | Adjuvanted RSVPreF3 | Placebo group | Adjuvanted RSVPreF3 | Placebo group |
|  | group (N=5014) | (N=4954) | group (N=7455) | (N=7549) |
| Mean age ± SD, years | 70.2 ± 6.6 | 70.2 ± 6.6 | 69.1 ± 6.4 | 69.1 ± 6.3 |
| Age category, n (%) |  |  |  |  |
| 60–69 years | 2582 (51.5) | 2558 (51.6) | 4380 (58.8) | 4424 (58.6) |
| 70–79 years | 1977 (39.4) | 1920 (38.8) | 2512 (33.7) | 2573 (34.1) |
| ≥80 years | 455 (9.1) | 476 (9.6) | 563 (7.6) | 552 (7.3) |
| Female sex, n (%) | 2357 (47.0) | 2376 (48.0) | 4132 (55.4) | 4053 (53.7) |
| Race, n (%) |  |  |  |  |
| African | 356 (7.1) | 364 (7.3) | 708 (9.5) | 737 (9.8) |
| Asian | 298 (5.9) | 288 (5.8) | 655 (8.8) | 668 (8.8) |
| White | 4151 (82.8) | 4112 (83.0) | 5738 (77.0) | 5824 (77.1) |
| Other | 209 (4.2) | 190 (3.8) | 354 (4.7) | 320 (4.2) |
| Hispanic or Latino ethnicity, n (%) | 285 (5.7) | 255 (5.1) | 397 (5.3) | 427 (5.7) |
| Mean body mass index ± SD, kg/m<sup>2</sup> | 30.5 ± 6.5 | 30.5 ± 6.2 | 28.2 ± 5.6 | 28.2 ± 5.7 |
| $\geq 1$ pre-existing condition of interest<sup>a</sup>, n (%) | 5014 (100) | 4954 (100) | 0 | 0 |
| $\geq 1$ cardiorespiratory condition of interest, n (%) | 2577 (51.4) | 2505 (50.6) | 0 | 0 |
| $\geq 1$ endocrine/metabolic condition of interest, n (%) | 3243 (64.7) | 3276 (66.1) | 0 | 0 |

Pre-existing conditions of interest were cardiorespiratory conditions (i.e., chronic respiratory or pulmonary disease [including chronic obstructive pulmonary disease and asthma] and chronic heart failure) and endocrine or metabolic conditions (i.e., type 1 or type 2 diabetes and advanced liver or renal disease).
Adjuvanted RSVPreF3 group, participants who received a dose of adjuvanted respiratory syncytial virus (RSV) prefusion F protein-based vaccine (adjuvanted RSVPreF3) pre-RSV season 1; placebo group, participants who received a dose of placebo pre-RSV season 1; N, number of participants in the exposed population; SD, standard deviation; n (%), number (percentage) of participants with the specified characteristic.
<sup>a</sup>More than one type of pre-existing condition of interest could occur in a single participant.

### Vaccine efficacy against RSV-LRTD and RSV-ARI in participants with pre-existing medical conditions

The median follow-up in the modified exposed population for the efficacy analysis of a single vaccine dose was 30.6 months (IQR: 26.2–32.0). The observed incidence rates (per 1000 person-years) in the placebo group of both RSV-LRTD and RSV-ARI were higher among participants with ≥1 pre-existing condition of interest (10.8 for RSV-LRTD and 18.0 for RSV-ARI) than among those without any of these conditions (6.0 and 14.4, respectively). This was especially pronounced for participants with COPD (16.5 and 23.3) and asthma (18.3 and 23.6) (**Figure 3**).

**Figure 3.**
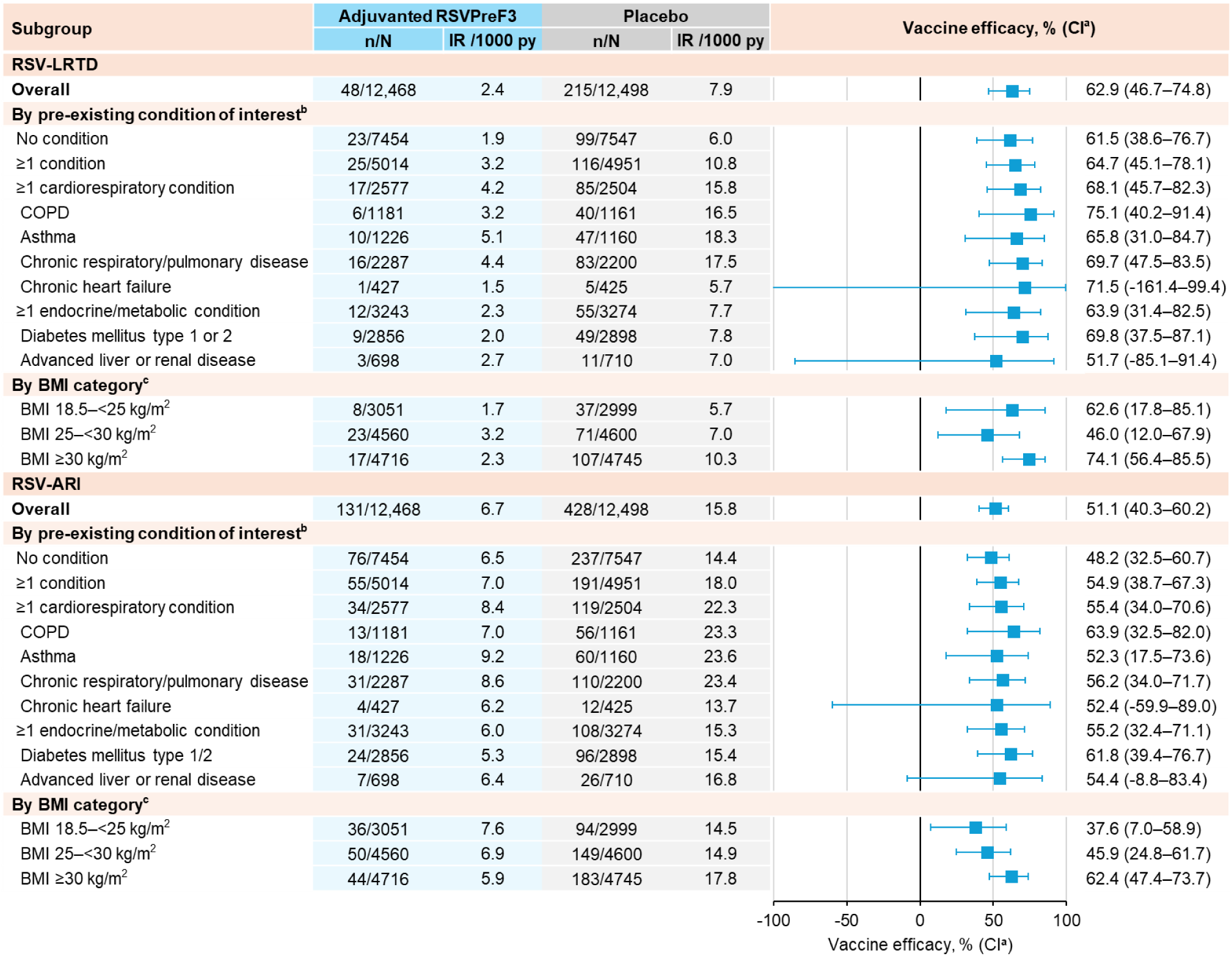
Vaccine efficacy of a single adjuvanted RSVPreF3 dose against first occurrence of RSV-LRTD and RSV-ARI over three RSV seasons, overall and among participants with pre-existing medical conditions (modified exposed population) Results based on post-hoc exploratory analyses, except for those in the overall trial population, in participants with no condition, ≥1 condition, ≥1 cardiorespiratory, and ≥1 endocrine/metabolic condition of interest. Analysis included data collected from day 15 post-dose 1 until end of Northern Hemisphere season 3 (efficacy data lock point: 30 April 2024) or until end of Southern Hemisphere season 1 (efficacy data lock point: 30 September 2022) for participants who did not receive dose 2. Participants who received adjuvanted RSVPreF3 as dose 1 and placebo as dose 2 (RSV single-dose group) and participants in the placebo group contributed to the entire follow-up. Participants who received adjuvanted RSVPreF3 as doses 1 and 2 (RSV revaccination group) contributed to season 1 but were censored at dose 2. Vaccine efficacy was estimated using a Poisson model adjusted for age, region, and season. Adjuvanted RSVPreF3, adjuvanted respiratory syncytial virus (RSV) prefusion F protein-based vaccine; RSV-LRTD, RSV-related lower respiratory tract disease confirmed by the adjudication committee; RSV-ARI, RSV-related acute respiratory illness; n, number of participants with ≥1 RSV-LRTD or RSV-ARI; N, number of participants in the modified exposed population; IR, incidence rate of participants reporting ≥1event, expressed per 1000 person-years (py), calculated as 1000 x (n/T), with T the sum of follow-up time from day 15 post-dose 1 until first occurrence of the event, data lock point, or drop-out; CI, confidence interval; COPD, chronic obstructive pulmonary disease; BMI, body mass index (18.5–<25 kg/m^2^: healthy weight, 25–<30 kg/m^2^: overweight, ≥30 kg/m^2^: obesity). ^a^97.5% CI for RSV-LRTD, overall (confirmatory secondary endpoint); 95% CI for other endpoints. ^b^More than one type of pre-existing condition of interest could occur in a single participant. ^c^Among the 131 (adjuvanted RSVPreF3) and 145 (placebo) participants with a BMI <18.5 kg/m^2^, none reported an RSV-LRTD, and one (adjuvanted RSVPreF3) and two (placebo) reported an RSV-ARI.

Among participants with ≥1 pre-existing condition of interest, the cumulative efficacy of a single vaccine dose over three RSV seasons was 64.7% (95% CI: 45.1–78.1) against first occurrence of RSV-LRTD, with 25 cases in the vaccine and 116 in the placebo group, 54.9% (38.7–67.3) against RSV-ARI, with 55 cases in the vaccine and 191 in the placebo group (**Figure 3**), and 70.7% (36.9–88.1) against severe RSV-LRTD, with 8 cases in the vaccine and 45 in the placebo group (**Supplementary Table 1**, post-hoc). Post-hoc analyses showed efficacy estimates of 75.1% (40.2–91.4; RSV-LRTD; 6 vs 40 cases) and 63.9% (32.5–82.0; RSV-ARI; 13 vs 56 cases) among participants with COPD, 65.8% (31.0–84.7; RSV-LRTD; 10 vs 47 cases) and 52.3% (17.5–73.6; RSV-ARI; 18 vs 60 cases) among those with asthma, 69.8% (37.5–87.1; RSV-LRTD; 9 vs 49 cases) and 61.8% (39.4–76.7; RSV-ARI; 24 vs 96 cases) among those with diabetes, and 74.1% (56.4–85.5; RSV-LRTD; 17 vs 107 cases) and 62.4% (47.4–73.7; RSV-ARI; 44 vs 183 cases) among those with obesity (BMI ≥30 kg/m^2^). The subgroups with chronic heart failure and advanced liver or renal disease were too small to reliably estimate vaccine efficacy (**Figure 3**). The efficacy of a single dose by season and of the revaccination regimen over three seasons are presented in the **Supplementary results** and **Supplementary Tables 2−5**.

### Vaccine efficacy against complications and disease exacerbation related to RSV-ARI

From day 15 post-dose 1 until the end of the third RSV season, 62 participants who received one vaccine dose or placebo reported complications related to RSV-ARI. Approximately two-thirds of these (43/62) were individuals with ≥1 pre-existing cardiorespiratory condition of interest (**Figure 4**). All reported complications were respiratory, except for otitis media in two participants in the placebo group and new-onset congestive heart failure in a participant in the placebo group who also reported exacerbation of asthma (**Supplementary Table 6**).

**Figure 4.**
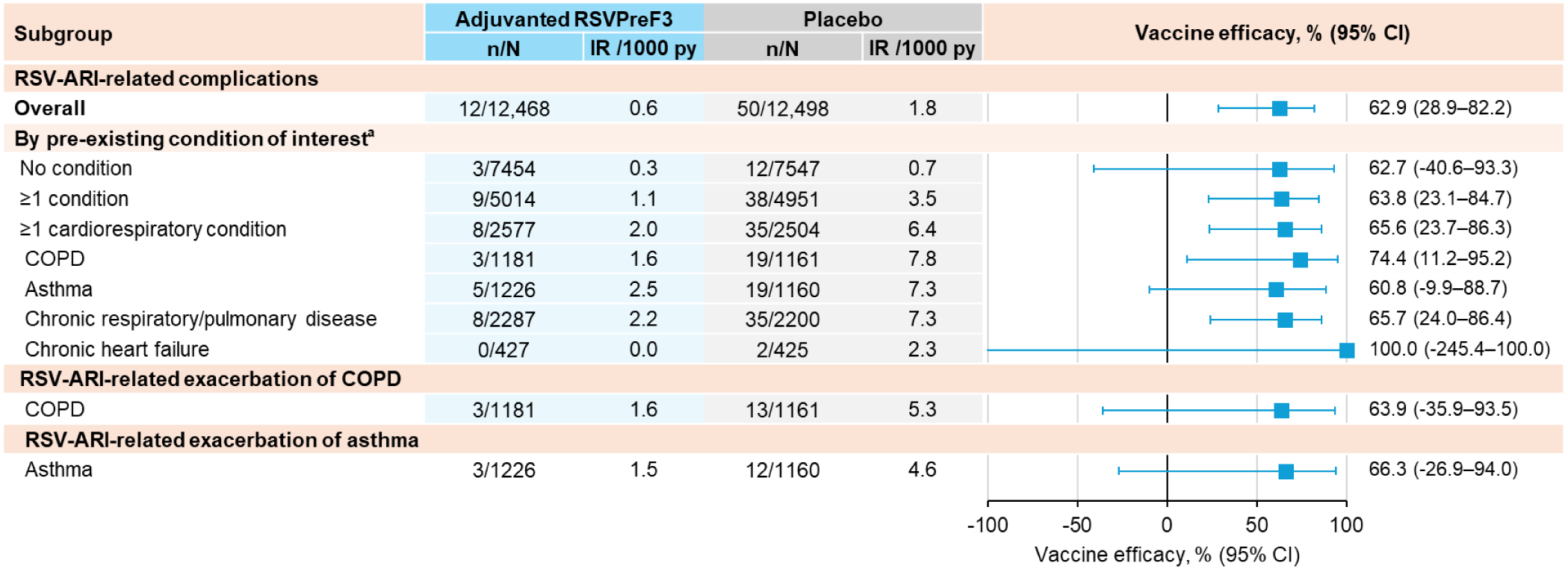
Vaccine efficacy of a single adjuvanted RSVPreF3 dose against RSV-ARI-related complications and disease exacerbations over three RSV seasons, overall and among participants with pre-existing medical conditions (modified exposed population) Results based on post-hoc exploratory analyses, except for those in the overall trial population. Complications could include respiratory complications (pneumonia, new diagnosis or exacerbation of COPD or asthma, other respiratory complications [e.g., sinusitis, bronchitis, dyspnoea]) and non-respiratory complications (new-onset or worsening congestive heart failure, myocardial infarction, stroke, diabetes, other non-respiratory complications). Analysis included data collected from day 15 post-dose 1 until end of Northern Hemisphere season 3 (efficacy data lock point: 30 April 2024) or until end of Southern Hemisphere season 1 (efficacy data lock point: 30 September 2022) for participants who did not receive dose 2. Participants who received adjuvanted RSVPreF3 as dose 1 and placebo as dose 2 (RSV single-dose group) and participants in the placebo group contributed to the entire follow-up. Participants who received adjuvanted RSVPreF3 as doses 1 and 2 (RSV revaccination group) contributed to season 1 but were censored at dose 2. Vaccine efficacy was estimated using a Poisson model adjusted for age, region, and season. Adjuvanted RSVPreF3, adjuvanted respiratory syncytial virus (RSV) prefusion F protein-based vaccine; RSV-ARI, RSV-related acute respiratory illness; n, number of participants with ≥1 RSV-ARI-related complication; N, number of participants in the modified exposed population; IR, incidence rate of participants reporting ≥1event, expressed per 1000 person-years (py), calculated as 1000 x (n/T), with T the sum of follow-up time from day 15 post-dose 1 until first occurrence of the event, data lock point, or drop-out; CI, confidence interval; COPD, chronic obstructive pulmonary disease; BMI, body mass index (18.5–<25 kg/m^2^: healthy weight, 25–<30 kg/m^2^: overweight, ≥30 kg/m^2^: obesity). ^a^More than one type of pre-existing condition of interest could occur in a single participant.

The cumulative efficacy over three RSV seasons against RSV-ARI-related complications was 62.9% (95% CI: 28.9–82.2) in the overall trial population, with 12 cases in the vaccine group vs 50 in the placebo group. In post-hoc analyses, efficacy was 65.6% (23.7–86.3; 8 vs 35 cases) among participants with ≥1 pre-existing cardiorespiratory condition of interest, 74.4% (11.2–95.2; 3 vs 19 cases) among those with COPD, and 60.8% (−9.9–88.7; 5 vs 19 cases) among those with asthma. Among participants with COPD, a cumulative efficacy of 63.9% (95% CI: −35.9–93.5; 3 vs 13 cases) was observed against RSV-ARI-related COPD exacerbations over the three RSV seasons. Among participants with asthma, an efficacy of 66.3% (−26.9–94.0; 3 vs 12 cases) was observed against RSV-ARI-related asthma exacerbations (**Figure 4**). Vaccine efficacy against complications during season 1 and over the first two seasons is presented in **Supplementary Table 7.**

### Medication use during RSV-ARI episodes

Post-hoc analyses showed that, from day 15 post-dose 1 until the end of season 3, 11/131 (8.4%) RSV-ARI episodes in participants who received a single vaccine dose and 42/435 (9.7%) RSV-ARI episodes in those who received placebo were treated with systemic corticosteroids. In addition, 26/131 (19.8%; adjuvanted RSVPreF3) and 124/435 (28.5%; placebo) RSV-ARI episodes were treated with antibiotics (**Table 2**). Treatment of RSV-ARI with systemic corticosteroids and antibiotics tended to be observed more frequently among participants with pre-existing cardiorespiratory conditions than in the overall study population and less frequently in vaccine than in placebo recipients (although CIs overlapped). Among participants with COPD, 2/13 (15.4%) RSV-ARI episodes in adjuvanted RSVPreF3 recipients and 13/58 (22.4%) in placebo recipients were treated with systemic corticosteroids, and 6/13 (46.2%; adjuvanted RSVPreF3) and 33/58 (56.9%; placebo) were treated with antibiotics (**Table 2**).

**Table 2.** Use of systemic corticosteroids and antibiotics to treat RSV-ARI episodes over three RSV seasons, overall and among participants with pre-existing medical conditions (modified exposed population)

| Medication<br>Subgroup | Adjuvanted RSVPreF3 group<br>(single dose) |  | Placebo group |  |
| --- | --- | --- | --- | --- |
|  | n/N | % (95% CI) | n/N | % (95% CI) |
| **Systemic corticosteroids** |  |  |  |  |
| Overall | 11/131 | 8.4 (4.3–14.5) | 42/435 | 9.7 (7.0–12.8) |
| ≥1 pre-existing condition of interest<sup>a</sup> | 7/55 | 12.7 (5.3–24.5) | 31/194 | 16.0 (11.1–21.9) |
| ≥1 cardiorespiratory condition<sup>a</sup> | 6/34 | 17.6 (6.8–34.5) | 26/122 | 21.3 (14.4–29.6) |
| COPD | 2/13 | 15.4 (1.9–45.4) | 13/58 | 22.4 (12.5–35.3) |
| Asthma | 4/18 | 22.2 (6.4–47.6) | 14/61 | 23.0 (13.2–35.5) |
| **Antibiotics** |  |  |  |  |
| Overall | 26/131 | 19.8 (13.4–27.7) | 124/435 | 28.5 (24.3–33.0) |
| ≥1 pre-existing condition of interest<sup>a</sup> | 16/55 | 29.1 (17.6–42.9) | 73/194 | 37.6 (30.8–44.9) |
| ≥1 cardiorespiratory condition<sup>a</sup> | 13/34 | 38.2 (22.2–56.4) | 53/122 | 43.4 (34.5–52.7) |
| COPD | 6/13 | 46.2 (19.2–74.9) | 33/58 | 56.9 (43.2–69.8) |
| Asthma | 6/18 | 33.3 (13.3–59.0) | 26/61 | 42.6 (30.0–55.9) |

Results based on post-hoc exploratory analyses. RSV-ARI, respiratory syncytial virus (RSV)-related acute respiratory illness; adjuvanted RSVPreF3, adjuvanted RSV prefusion F protein-based vaccine; n, number of RSV-ARI episodes treated with the indicated medication; N, number of RSV-ARI episodes reported from day 15 post-dose 1 until end of Northern Hemisphere season 3 (efficacy data lock point: 30 April 2024) or until end of Southern Hemisphere season 1 (efficacy data lock point: 30 September 2022) for participants who did not receive dose 2. Note, participants who received adjuvanted RSVPreF3 as doses 1 and 2 (RSV revaccination group) contributed to season 1 but were censored at dose 2; %, percentage of RSV-ARI episodes treated with the indicated medication (calculated as $100 \times [n/N]$ ); CI, confidence interval; COPD, chronic obstructive pulmonary disease.
<sup>a</sup>More than one type of pre-existing condition of interest could occur in a single participant.

Immune responses in participants with pre-existing medical conditions are presented in the **Supplementary results** and **Supplementary Figure 1**.

## Discussion

Previously published results from pre-specified analyses of the AReSVi-006 trial showed that a single dose of adjuvanted RSVPreF3 was efficacious in preventing RSV-LRTD and RSV-ARI over three consecutive RSV seasons among medically stable adults aged ≥60 years with ≥1 pre-existing medical condition associated with an increased risk of severe RSV disease [21]. The current post-hoc analyses suggest that the vaccine was also efficacious against RSV-LRTD and RSV-ARI among individual subgroups with COPD, asthma, diabetes, or obesity. In addition, the results suggest that adjuvanted RSVPreF3 might help prevent RSV-ARI-related complications in participants with COPD and asthma, including disease exacerbations. Furthermore, the observed proportion of RSV-ARI episodes that required systemic corticosteroids or antibiotics among participants with underlying cardiorespiratory conditions tended to be lower in vaccine than in placebo recipients.

Most older adults hospitalised with RSV have underlying medical conditions. A study based on surveillance data from the RSV-Associated Hospitalisation Surveillance Network in the United States found that 37.8% of adults aged ≥60 years hospitalised with RSV had obesity, 33.7% had COPD, 33.2% had congestive heart failure, and 32.6% had diabetes mellitus [28]. Several of these conditions have been shown to increase the risk of severe RSV disease and various complications [1–7]. Consistent with this, in our trial, we observed a higher incidence of RSV-ARI, RSV-LRTD, and RSV-ARI-related complications in participants with ≥1 pre-existing condition of interest, most notably COPD or asthma, than in those without any of the pre-existing conditions of interest. Given the high RSV burden in people with respiratory and metabolic conditions and the observed vaccine efficacy in our trial, these populations may stand to benefit most from RSV vaccination, and adjuvanted RSVPreF3 could have a considerable impact on public health.

RSV has been recognised as an important viral trigger of exacerbations of COPD and asthma [11,29,30]. Exacerbations impose a substantial burden on patients and healthcare systems and are associated with disease progression, accelerated loss of lung function and quality of life, increased hospitalisation rates, and greater mortality [15,31,32]. Preventing or minimising the risk of exacerbations is an important goal in the management of COPD and asthma to attain stable disease or remission [13,14,33]. To this end, the Global Initiative for Asthma advises healthcare professionals to encourage individuals with asthma to follow their local immunisation schedule, including for RSV [14]. Similarly, the Global Initiative for Chronic Obstructive Lung Disease states that people with COPD should receive all recommended vaccinations in line with local guidelines, including RSV vaccination [13]. The results of our post-hoc analyses support these recommendations; the observed incidence rates of RSV-ARI-related COPD and asthma exacerbations tended to be lower in the vaccine compared to the placebo group, with efficacy estimates of 63.9% (95% CI: −35.9–93.5) and 66.3% (−26.9–94.0), respectively. These results must be interpreted with caution due to the exploratory nature of the analyses and the low number of exacerbation events, which resulted in wide CIs including 0. Consistent with our results, a recent study in the United States found that adjuvanted RSVPreF3 was effective in preventing RSV-related severe chronic COPD exacerbations, severe asthma exacerbations, and major adverse cardiovascular events among ≥60-year-olds with underlying COPD, asthma, and cardiovascular disease, respectively [34].

Systemic corticosteroids are commonly used to treat exacerbations of COPD and asthma in adults [35,36], including those triggered by viral infections. However, long-term and repeated short-term use of systemic corticosteroids are associated with an increased risk of acute and chronic adverse events [35–38]. Moreover, a retrospective cohort study of adults admitted to the hospital with RSV showed no improvement in clinical outcomes with systemic corticosteroid treatment. Instead, an increase in the rate of bacterial infections and a longer duration of hospitalisation were found [10]. The results of our post-hoc analyses suggest a trend for a lower proportion of RSV-ARI episodes treated with systemic corticosteroids in patients with COPD vaccinated with adjuvanted RSVPreF3 (15.4%) compared to those who received placebo (22.4%), which is consistent with the lower incidence of COPD exacerbations observed in the vaccinated group. Antimicrobial resistance, driven by the inappropriate use and overuse of antibiotics, is a major threat to global health. Vaccines are an integral component of the multifaceted approach to contain antimicrobial resistance by preventing infections, thereby reducing antimicrobial use [39]. In our study, we saw a trend toward a reduction in the frequency of RSV-ARI episodes treated with antibiotics among vaccine vs placebo recipients with COPD (46.2% vs 56.9%) or asthma (33.3% vs 42.6%). However, the study was not designed to evaluate the impact of the vaccine on corticosteroid or antibiotic consumption, and CIs in the two groups overlapped. The lower frequency of corticosteroid and antibiotics use may suggest that the investigators perceived the RSV-ARI cases among vaccinated participants as less severe or requiring less medical intervention; however, other factors may contribute, and this should be interpreted cautiously. Patient-reported outcomes evaluated in the AReSVi-006 trial similarly indicated that RSV-associated symptoms in breakthrough RSV-ARI in vaccinated participants were less severe than those in RSV-ARI in unvaccinated participants [27].

Our post-hoc analyses suggest that a single dose of adjuvanted RSVPreF3 is efficacious against RSV-LRTD and RSV-ARI in participants with diabetes and in those with obesity. Several studies have shown significantly higher hospitalisation rates in RSV-infected adults with diabetes than in those without [1,2,4,5]. In addition, in adults aged ≥60 years hospitalised with RSV, diabetes was found to be associated with a persistent functional decline at 6 months post-discharge [40]. Although obesity has not consistently been established as an independent risk factor for severe RSV disease [2], a population-based cohort study in Spain did identify severe obesity (BMI ≥40 kg/m^2^) as an independent risk factor for RSV hospitalisation [5]. The United States Centers for Disease Control and Prevention also list BMI ≥40 kg/m^2^ as a risk factor for severe RSV disease among adults aged 60–74 years [41]. In addition, obesity is associated with several other chronic conditions, including cardiovascular disease and diabetes, and has been shown to increase the risk of severe outcomes and hospitalisation from various other respiratory viruses [42,43]. In our trial, the observed vaccine efficacy in participants with obesity may be influenced by coexisting comorbidities rather than obesity alone.

The analyses presented here have limitations. They were exploratory, and most were performed post hoc. Some of the subgroups were small, and no adjustment for multiplicity was done. Conclusions are therefore hypothesis-generating and should be interpreted with caution. In addition, no objective criteria were defined to capture exacerbations and complications, and their relationship to RSV-ARI was based on investigator assessment. This creates a risk of misclassification and differential assessment. Medication use outcomes are prone to confounding as the prescription of corticosteroids and antibiotics is clinician-dependent, region-dependent, and influenced by perceived severity, comorbid status, access to care, local antimicrobial stewardship, and diagnostic uncertainty. Another limitation is that our study only enrolled medically stable participants, and the results may thus not be entirely generalisable to those with serious or unstable chronic illnesses or with frequently exacerbating COPD or asthma. Finally, data on the severity of asthma and COPD exacerbations were not collected, nor were clinical characteristics, such as history of exacerbations, lung function, or symptom scores. The different subgroups may therefore represent heterogeneous groups of patients.

In summary, our findings from the post-hoc analyses presented here, together with the previously reported safety profile, based on approximately 3 years of follow-up after vaccination, suggest that the adjuvanted RSVPreF3 could improve public health by preventing RSV-LRTD and other respiratory complications from RSV in medically stable older adults with underlying medical conditions, like COPD and asthma. Our findings are supported by recent real-world-evidence from Denmark and the United States showing effectiveness of adjuvanted RSVPreF3 against RSV-related hospitalisation and complications among adults aged ≥60 years with asthma, COPD, cardiovascular disease, and diabetes [34,44,45]. The observed trend for a reduced use of systemic corticosteroids and antibiotics in breakthrough RSV infections among vaccinated participants warrants further investigation and, if confirmed, could indicate that RSV vaccination may mitigate adverse events and contribute to combatting antimicrobial resistance.

## Notes

## Acknowledgements

We are grateful to all participants; and to all the AReSVi-006 study group members, their staff, and their institutions. We also thank Akkodis Belgium for medical writing (by Natalie Denef), design, and publication coordination support on behalf of GSK.

## AReSVi-006 study group members (listed alphabetically)

Adams, Mark; Adams, Michael; Agutu, Clara; Akite, Elaine Jacqueline; Alt, Ingrid; Andrews, Charles; Asatryan, Asmik; Athan, Eugene; Bahrami, Ghazaleh; Bargagli, Elena; Bhorat, Qasim; Bird, Paul; Borowy, Przemyslaw; Boutry, Céline; Brotons Cuixart, Carles; Browder, David; Brown, Judith; Buntinx, Erik; Cameron, Donald; Cartier, Cyrille; Chinsky, Kenneth; Choi, Melissa; Choo, Eun-Ju; Collete, Delphine; Corral Carrillo, Maria; Cuadripani, Susanna; Davis, Matthew G; de Heusch, Magali; de Looze, Ferdinandus; De Meulemeester, Marc; De Negri, Ferdinando; DeAtkine, David; Dedkova, Viktoriya; Deraedt, Quentin; Descamps, Dominique; Dezutter, Nancy; Dzongowski, Peter; Eckermann, Tamara; Essink, Brandon; Faulkner, Karen; Ferguson, Murdo; Fuller, Gregory; Galan Melendez, Isabel Maria; Gentile, Ivan; Gerard, Catherine; Ghesquiere, Wayne; Grimard, Doria; Gruselle, Olivier; Halperin, Scott; Heer, Amardeep; Helman, Laura; Hotermans, Andre; Jelinek, Tomas; Kamerbeek, Jackie; Kim, Hyo Youl; Kimmel, Murray; Koch, Mark; Kokko, Satu; Koski, Susanna; Kotb, Shady; Lalueza, Antonio; Langley, Joanne M; Lee, Jin-Soo; Leroux-Roels, Isabel; Lins, Muriel; Lombaard, Johannes; Mahomed, Akbar; Malerba, Mario; Marion, Sandie; Martinon-Torres, Federico; Martinot, Jean-Benoit; Masuet-Aumatell, Cristina; McNally, Damien; Medina Pech, Carlos Eduardo; Mendez Galvan, Jorge; Mercati, Lise; Mesotten, Dieter; Mitha, Essack; Mngadi, Kathryn; Moeckesch, Beate; Montgomery, Barnaby; Murray, Linda; Nally, Rhiannon; Narejos Perez, Silvia; Newberg, Joseph; Nugent, Paul; Ochoa Mazarro, Dolores; Oda, Harunori; Orso, Maurizio; Ortiz Molina, Jacinto; Pak, Tatiana; Park, Dae Won; Patel, Meenakshi; Patel, Minesh; Pedro Pijoan, Anna Maria; Perez, Alberto Borobia; Perez-Breva, Lina; Perez Vera, Merce; Pileggi, Claudia; Pregliasco, Fabrizio; Pretswell, Carol; Quinn, Dean; Reynolds, Michele; Romanenko, Viktor; Rosen, Jeffrey; Roy, Nathalie; Ruiz Antoran, Belen; Sakata, Hideaki; Sauter, Joachim; Schaefer, Axel; Sein Anand, Izabela; Serra Rexach, Jose Antonio; Shu, David; Siig, Andres; Simon, William; Smakotina, Svetlana; Steenackers, Katie; Stephan, Brigitte; Tafuri, Silvio; Takazawa, Kenji; Tellier, Guy; Terryn, Wim; Tharenos, Leslie; Thomas, Nick; Toursarkissian, Nicole; Ukkonen, Benita; Vale, Noah; Van Landegem, Pieter-Jan; van Zyl-Smith, Richard N; Vanden Abeele, Carline; Verheust, Céline; Vermeersch, Lode; Vitale, Francesco; Voloshyna, Olga; White, Judith; Wie, Seong-Heon; Wilson, Jonathan; Ylisastigui, Pedro; Zocco, Manuel.

## Funding

This work was supported by GSK who funded this trial, was involved in all stages of the study design, conduct, and analysis, and took charge of all costs associated with the development and publication of this manuscript.

## Competing interests

**A Papi** declares funding from GSK paid to his institution for conducting the trial; grants from GSK, Chiesi, AstraZeneca, and Sanofi; consulting fees and/or honoraria from GSK, Chiesi, AstraZeneca, Sanofi, Avillion, Moderna, Roche, Regeneron, Zambon, and Zentiva; **DMG Halpin** declares honoraria from AstraZeneca, Boehringer Ingelheim, Chiesi, GSK, Inogen, Menarini, Novartis, Pfizer, and Sanofi; payment from Chiesi and Synairgen for participating on data safety monitoring boards or advisory boards. **RG Feldman** declares payments from GSK for expert testimony and lectures. **MG Ison** declares that research support from GSK was paid to his previous institution, Northwestern University; he received consulting fees paid by Adagio Therapeutics, ADMA Biologics, Adamis Pharmaceuticals, AlloVir, Atea, Cidara Therapeutics, Genentech/Roche, Janssen, Shionogi, Takeda, Talaris, and Eurofins Viracor; and payment for participation in data safety monitoring boards or advisory boards from Adamis Pharmaceuticals, AlloVir, National Institutes of Health, CSL Behring, Janssen, Merck, Seqirus, Takeda, and Talaris; all of these ended in December, 2022. **MG Ison** also receives author royalties from UpToDate, which is ongoing, and serves as Chair of the International Society for Influenza and other Respiratory Virus Diseases Antiviral Group (separate from his government service), and used to be Editor-in-Chief of *Transplant Infectious Disease*. **MG Ison** was a consultant for Romark but received no consulting fees. **TF Schwarz** reports honoraria from AstraZeneca, Bavarian Nordic, Biogen, BioNTech, CSL-Seqirus, CSL-Vifor, Diasorin, GSK, Janssen-Cilag, Merck-Serono, Moderna, MSD, Novavax, Pfizer, Roche, Sanofi-Aventis, Synlab, and Takeda; and participation on advisory boards from Bavarian Nordic, BioNTech, CSL-Seqirus, GSK, Moderna, Novavax, and Takeda. **R Antonelli Incalzi** declares that his institution received a grant from GSK for conducting the trial. **L Fissette**, **S Xavier**, **M-P David**, **J-P Michaud**, **S Kotb**, **C Maréchal**, **A Olivier**, **V Hulstrøm**, and **M Van der Wielen** are employed by GSK; **L Fissette**, **M-P David**, **J-P Michaud**, **S Kotb**, **C Maréchal**, **A Olivier**, **V Hulstrøm**, and **M Van der Wielen** hold financial equities in GSK. **M Van der Wielen** has stock options from Haleon. **L Fissette**, **M-P David**, and **M Van der Wielen** are part of vaccine patents filed by GSK. **D-G Lee** declares no competing interests. The authors declare no other financial or non-financial relationships.

**MG Ison** is currently employed by the National Institutes of Health, but his role in the trial started prior to this employment. The content of this publication is solely the responsibility of the authors and does not necessarily represent the official views of the National Institutes of Health.

## Author contributions

**L Fissette**, **M-P David**, **J-P Michaud**, **S Kotb**, **C Maréchal**, **A Olivier**, **V Hulstrøm**, and **M Van der Wielen** were involved in the conception or design of the study. **A Papi**, **RG Feldman**, **MG Ison**, **TF Schwarz**, **D-G Lee**, and **R Antonelli Incalzi** participated in the acquisition or generation of the data. **L Fissette**, **S Xavier**, and **M-P David** performed the data analysis. **A Papi**, **DMG Halpin**, **RG Feldman**, **MG Ison**, **TF Schwarz**, **D-G Lee**, **R Antonelli Incalzi**, **L Fissette**, **S Xavier**, **M-P David**, **J-P Michaud**, **S Kotb**, **C Maréchal**, **A Olivier**, **V Hulstrøm**, and **M Van der Wielen** were involved in the interpretation of the data. All authors had access to the data, critically revised the manuscript drafts, approved the final submitted version, and were responsible for the decision to submit this manuscript for publication.

## Data availability

Please refer to the link https://www.gsk-studyregister.com/en/ to access GSK’s data sharing policies and to request anonymised participant-level data.

## Trademark statement/other disclaimers

Arexvy and AS01 are trademarks licensed to or owned by GSK. AS01_E_ is an adjuvant system containing 25 µg 3-O-desacyl-4′-monophosphoryl lipid A, 25 µg QS-21, and liposome (QS-21 is licensed by GSK from Antigenics LLC, a wholly owned subsidiary of Agenus Inc., a Delaware, USA corporation).

## Notes

### Clinical Trial

NCT04886596

### Clinical Protocols

https://www.gsk-studyregister.com/en/trial-details/?id=212494

### Author Declarations

Internal approval was obtained from GSK to conduct the study. Appropriate ethical oversight was obtained from corresponding IRBs/EC for all participating sites.

