## Supplement for "Adjuvanted RSVPreF3 vaccine impact over 3 RSV seasons in older adults with comorbidities"

### Supplementary methods

#### *Symptoms and signs used to define acute respiratory illness (ARI) and lower respiratory tract disease (LRTD)*

Respiratory symptoms were: nasal congestion/rhinorrhoea; sore throat; new or increased sputum (lower respiratory symptom); new or increased cough (lower respiratory symptom); new or increased dyspnoea (shortness of breath) (lower respiratory symptom).

Respiratory signs were: new or increased wheezing (reported by the participant or investigator) (lower respiratory sign); new or increased crackles/rhonchi (reported by the investigator) based on chest auscultation (lower respiratory sign); respiratory rate ≥20 respirations/min (reported by the investigator) (lower respiratory sign); low or decreased oxygen saturation (= oxygen saturation <95% or ≤90% if pre-season baseline was <95%, reported by the investigator) (lower respiratory sign); need for oxygen supplementation (reported by the investigator) (lower respiratory sign).

Systemic symptoms/signs were: fever (body temperature ≥38.0°C/100.4°F by any route) or feverishness (feeling of having fever without objective measurement); fatigue; body aches; headache; decreased appetite.

An LRTD was classified as severe (1) if the episode was assessed as severe by the investigator or if the participant experienced at least two lower respiratory signs (as listed above), or (2) if the participant required oxygen supplementation, positive airway pressure therapy, or other types of mechanical ventilation, or any significant change or adaptation in one of these therapies if already used [1].

#### *Complications*

The following complications were collected throughout the study, starting on day 1.

<u>Respiratory complications:</u>

- Pneumonia: a clinical diagnosis of pneumonia based on signs and symptoms, with or without chest radiograph that demonstrated a new or progressive infiltrate.
- New diagnosis of chronic obstructive pulmonary disease (COPD) or exacerbation of COPD: a new diagnosis of COPD or, in a participant with previously diagnosed COPD, a worsening of COPD necessitating an increase in the dosage or modification of the administered treatment.
- New diagnosis of asthma or exacerbation of asthma: a new diagnosis of asthma or, in a participant with previously diagnosed asthma, a worsening of asthma necessitating an increase in the dosage or modification of the administered treatment.
- Other respiratory complications: other diagnosis of respiratory illness, including new diagnosis or exacerbation of a pre-existing respiratory disease (e.g., emphysema, chronic bronchitis).

Non-respiratory complications:

- New onset or worsening of congestive heart failure
- Myocardial infarction
- Stroke
- Diabetes
- Other non-respiratory complications judged related to an ARI episode by the investigator

***Immunogenicity assessments***

Respiratory syncytial virus (RSV)-A and RSV-B neutralising titres were measured (as described previously [1]) in a subset of participants from a selected number of countries and trial sites (immunogenicity cohort) using serum samples collected on day 1 (prior to dose 1), day 31, pre-season 2 (prior to dose 2), and pre-season 3.

Immunogenicity was evaluated in the per-protocol population (protocol-compliant participants in the immunogenicity cohort who had post-dose immunogenicity data available). RSV-A and RSV-B neutralising geometric mean titres (GMTs) and mean geometric increases (MGIs) were calculated with 95% confidence intervals in subgroups of participants without any of the pre-existing conditions of interest, with ≥1 condition, ≥1 cardiorespiratory, and ≥1 endocrine or metabolic condition.

### Supplementary results

#### ***Efficacy of revaccination regimen against RSV-LRTD and RSV-ARI***

The cumulative efficacy of the revaccination regimen (two doses given approximately 1 year apart) over three RSV seasons post-dose 1 was generally in the same range as that of a single vaccine dose among the different subgroups of participants with pre-existing medical conditions, although point estimates for the revaccination regimen were numerically higher for most subgroups (**Supplementary Table 5**).

#### ***Immune response and persistence in participants with pre-existing medical conditions***

Regardless of the presence of pre-existing medical conditions of interest, RSV-A and RSV-B neutralising GMTs increased from pre-vaccination to day 31, with MGIs versus pre-vaccination ranging from 9.4 to 11.7 for RSV-A and 8.1 to 9.3 for RSV-B across groups (RSV revaccination and RSV single-dose) and subgroups (no condition of interest, ≥1 condition, ≥1 cardiorespiratory condition, ≥1 endocrinometabolic condition) (**Supplementary Figure 1**). Neutralising GMTs decreased over time but remained above baseline levels until at least pre-RSV season 3 irrespective of the vaccination schedule and the presence of pre-existing conditions, with MGIs versus pre-vaccination ranging from 2.2 to 3.5 for RSV-A and 1.6 to 2.4 for RSV-B across groups and subgroups (**Supplementary Figure 1**).

**Supplementary Table 1. Vaccine efficacy of a single adjuvanted RSVPreF3 dose against first occurrence of severe RSV-LRTD over three RSV seasons, overall and among participants with pre-existing medical conditions (modified exposed population)**

| Endpoint | Adjuvanted RSVPreF3 group | | | | Placebo group | | | | Vaccine efficacy, |
|---|---|---|---|---|---|---|---|---|---|
| **Subgroup** | **N** | **n** | **T, py** | **IR per 1000 py** | **N** | **n** | **T, py** | **IR per 1000 py** | **% (95% CI)** |
| **Severe RSV-LRTD**[a] | | | | | | | | | |
| Overall | 12,468 | 15 | 19,778.4 | 0.8 | 12,498 | 75 | 27,526.4 | 2·7 | 67.4 (42.4–82.7) |
| By pre-existing condition of interest[b] | | | | | | | | | |
| No condition | 7454 | 7 | 11,828.1 | 0.6 | 7547 | 30 | 16,711.0 | 1.8 | 63.5 (14.3–86.6) |
| ≥1 condition | 5014 | 8 | 7950.3 | 1.0 | 4951 | 45 | 10,815.4 | 4.2 | 70.7 (36.9–88.1) |
| ≥1 cardiorespiratory condition | 2577 | 6 | 4059.0 | 1.5 | 2504 | 39 | 5441.5 | 7.2 | 73.8 (37.4–91.0) |
| ≥1 endocrine/metabolic condition | 3243 | 4 | 5159.6 | 0.8 | 3274 | 15 | 7189.2 | 2.1 | 57.2 (−36.2–89.8) |

Results based on post-hoc exploratory analyses, except for those in the overall trial population. Analysis included data collected from day 15 post-dose 1 until end of Northern Hemisphere season 3 (efficacy data lock point: 30 April 2024) or until end of Southern Hemisphere season 1 (efficacy data lock point: 30 September 2022) for participants who did not receive dose 2. Participants who received adjuvanted RSVPreF3 as dose 1 and placebo as dose 2 (RSV single-dose group) and participants in the placebo group contributed to the entire follow-up. Participants who received adjuvanted RSVPreF3 as doses 1 and 2 (RSV revaccination group) contributed to season 1 but were censored at dose 2. Vaccine efficacy was estimated using a Poisson model adjusted for age, region, and season.

Adjuvanted RSVPreF3, adjuvanted respiratory syncytial virus (RSV) prefusion F protein-based vaccine; RSV-LRTD, RSV-related lower respiratory tract disease confirmed by the adjudication committee; N, number of participants in the modified exposed population; n, number of participants with ≥1 severe RSV-LRTD; T, sum of follow-up time from day 15 post-dose 1 until first occurrence of the event, data lock point, or drop-out; IR, incidence rate of participants reporting ≥1event, expressed per 1000 person-years (py), calculated as 1000 x (n/T); CI, confidence interval.

[a]Severe disease based on clinical signs or investigator assessment or on the need for supportive therapy. [b]More than one type of pre-existing condition of interest could occur in a single participant.

**Supplementary Table 2. Vaccine efficacy of a single adjuvanted RSVPreF3 dose against first occurrence of RSV-LRTD and RSV-ARI during the first RSV season, overall and among participants with pre-existing medical conditions (modified exposed population)**

| Endpoint | Adjuvanted RSVPreF3 group | | | | Placebo group | | | | Vaccine efficacy, |
|---|---|---|---|---|---|---|---|---|---|
| Subgroup | N | n | T, py | IR per 1000 py | N | n | T, py | IR per 1000 py | % (CI[a]) |
| **RSV-LRTD** | | | | | | | | | |
| Overall | 12,466 | 7 | 6865.9 | 1.0 | 12,494 | 40 | 6857.3 | 5.8 | 82.6 (57.9–94.1) |
| By pre-existing condition of interest[b] | | | | | | | | | |
| COPD | 1131 | 1 | 644.9 | 1.6 | 1113 | 8 | 619.6 | 12.9 | 88.1 (11.2–99.73) |
| Asthma | 1193 | 0 | 679.3 | 0.0 | 1112 | 4 | 629.2 | 6.4 | 100 (-5.5–100) |
| Chronic respiratory/pulmonary disease | 2223 | 1 | 1260.6 | 0.8 | 2122 | 12 | 1190.2 | 10.1 | 92.2 (47.3–99.8) |
| Chronic heart failure | 398 | 0 | 219.6 | 0.0 | 403 | 2 | 220.5 | 9.1 | 100 (-264.0–100) |
| Diabetes mellitus type 1 or 2 | 2829 | 0 | 1589.8 | 0.0 | 2875 | 12 | 1603.5 | 7.5 | 100 (71.4–100) |
| Advanced liver or renal disease | 667 | 0 | 370.7 | 0.0 | 674 | 2 | 380.0 | 5.3 | 100 (-257.2–100) |
| By BMI category[c] | | | | | | | | | |
| BMI 18.5–<25 kg/m$^2$ | 3051 | 2 | 1638.5 | 1.2 | 2997 | 5 | 1607.1 | 3.1 | 60.3 (-143.1–96.2) |
| BMI 25–<30 kg/m$^2$ | 4556 | 5 | 2511.2 | 2.0 | 4599 | 13 | 2521.6 | 5.2 | 61.3 (-15.7–89.2) |
| BMI ≥30 kg/m$^2$ | 4718 | 0 | 2648.7 | 0.0 | 4744 | 22 | 2655.7 | 8.3 | 100 (85.5–100) |

| | | | | | | | | | |
|---|---|---|---|---|---|---|---|---|---|
| **RSV-ARI** | | | | | | | | | |
| Overall | 12,466 | 27 | 6858.7 | 3.9 | 12,494 | 95 | 6837.8 | 13.9 | 71.7 (56.2–82.3) |
| By pre-existing condition of interest[b] | | | | | | | | | |
| COPD | 1131 | 1 | 644.9 | 1.6 | 1113 | 16 | 616.9 | 25.9 | 94.0 (61.4–99.9) |
| Asthma | 1193 | 1 | 678.8 | 1.5 | 1112 | 8 | 628.0 | 12.7 | 88.2 (11.9–99.7) |
| Chronic respiratory/pulmonary disease | 2223 | 2 | 1260.0 | 1.6 | 2122 | 22 | 1186.9 | 18.5 | 91.4 (65.0–99.0) |
| Chronic heart failure | 398 | 1 | 219.1 | 4.6 | 403 | 4 | 219.8 | 18.2 | 75.2 (-150.6–99.5) |
| Diabetes mellitus type 1 or 2 | 2829 | 6 | 1587.3 | 3.8 | 2875 | 28 | 1598.2 | 17.5 | 78.4 (46.9–92.7) |
| Advanced liver or renal disease | 667 | 0 | 370.7 | 0.0 | 674 | 5 | 378.8 | 13.2 | 100 (19.5–100) |
| By BMI category[c] | | | | | | | | | |
| BMI 18.5–<25 kg/m$^2$ | 3051 | 8 | 1636.3 | 4.9 | 2997 | 19 | 1602.0 | 11.9 | 58.1 (-0.2–84.2) |
| BMI 25–<30 kg/m$^2$ | 4556 | 13 | 2508.4 | 5.2 | 4599 | 29 | 2516.1 | 11.5 | 54.8 (10.3–78.5) |
| BMI ≥30 kg/m$^2$ | 4718 | 5 | 2646.5 | 1.9 | 4744 | 47 | 2646.8 | 17.8 | 89.4 (73.5–96.7) |

Results based on post-hoc exploratory analyses, except for those in the overall trial population. Analysis included cases collected from day 15 post-dose 1 until end of Northern Hemisphere season 1 (efficacy data lock point: 11 April 2022). Vaccine efficacy was estimated using a Poisson model adjusted for age and region.

Adjuvanted RSVPreF3, adjuvanted respiratory syncytial virus (RSV) prefusion F protein-based vaccine; RSV-LRTD, RSV-related lower respiratory tract disease confirmed by the adjudication committee; RSV-ARI, RSV-related acute respiratory illness; N, number of participants in the modified exposed population; n, number of participants with ≥1 RSV-LRTD or RSV-ARI; T, sum of follow-up time from day 15 post-dose 1 until first occurrence of the event, data lock point, or drop-out; IR, incidence rate of participants reporting at least one event, expressed per 1000 person-years (py), calculated as 1000 x (n/T); CI, confidence interval; COPD, chronic obstructive pulmonary disease; BMI, body mass index (18.5–<25 kg/$m^2$: healthy weight; 25–<30 kg/$m^2$: overweight; ≥30 kg/$m^2$: obesity).

[a]96.95% CI for RSV-LRTD, overall (confirmatory secondary endpoint); 95% CI for other endpoints. [b]More than one type of pre-existing condition of interest could occur in a single participant. [c]Among the 131 (adjuvanted RSVPreF3) and 145 (placebo) participants with a BMI <18.5 kg/$m^2$, none reported an RSV-LRTD and one (adjuvanted RSVPreF3) reported an RSV-ARI.

**Supplementary Table 3. Vaccine efficacy of a single adjuvanted RSVPreF3 dose against first occurrence of RSV-LRTD and RSV-ARI during the second RSV season, overall and among participants with pre-existing medical conditions (dose 2 modified exposed population)**

| Endpoint | Adjuvanted RSVPreF3 group (single dose) | | | | Placebo group | | | | Vaccine efficacy, % |
|---|---|---|---|---|---|---|---|---|---|
| Subgroup | N | n | T, py | IR per 1000 py | N | n | T, py | IR per 1000 py | (95% CI) |
| **RSV-LRTD** | | | | | | | | | |
| Overall | 4991 | 20 | 2448.8 | 8.2 | 10,031 | 91 | 4914.5 | 18.5 | 56.1 (28.2–74.4) |
| By pre-existing condition of interest[a] | | | | | | | | | |
| COPD | 454 | 1 | 231.7 | 4.3 | 868 | 17 | 431.7 | 39.4 | 88.3 (25.0–99.7) |
| Asthma | 485 | 6 | 237.7 | 25.2 | 929 | 24 | 455.8 | 52.7 | 53.0 (-18.2–84.3) |
| Chronic respiratory/pulmonary disease | 889 | 7 | 442.6 | 15.8 | 1720 | 38 | 850.6 | 44.7 | 66.0 (22.8–87.2) |
| Chronic heart failure | 162 | 0 | 81.9 | 0.0 | 312 | 2 | 159.7 | 12.5 | 100 (-438.1–100) |
| Diabetes mellitus type 1 or 2 | 1141 | 6 | 570.2 | 10.5 | 2295 | 16 | 1146.9 | 14.0 | 25.7 (-99.9–76.2) |
| Advanced liver or renal disease | 284 | 1 | 147.3 | 6.8 | 560 | 3 | 290.0 | 10.3 | 36.5 (-698.1–98.8) |
| By BMI category[b] | | | | | | | | | |
| BMI 18.5–<25 kg/m$^2$ | 1193 | 3 | 582.8 | 5.1 | 2402 | 17 | 1179.0 | 14.4 | 63.3 (-27.2–93.1) |
| BMI 25–<30 kg/m$^2$ | 1842 | 10 | 918.7 | 10.9 | 3705 | 31 | 1837.8 | 16.9 | 36.3 (-33.5–72.1) |
| BMI ≥30 kg/m$^2$ | 1901 | 7 | 923.9 | 7.6 | 3809 | 43 | 1848.3 | 23.3 | 68.2 (28.6–87.9) |

| | | | | | | | | | |
|---|---|---|---|---|---|---|---|---|---|
| **RSV-ARI** | | | | | | | | | |
| Overall | 4991 | 54 | 2438.1 | 22.1 | 10,031 | 181 | 4888.0 | 37.0 | 40.6 (19.0–57.0) |
| By pre-existing condition of interest[a] | | | | | | | | | |
| COPD | 454 | 4 | 230.5 | 17.4 | 868 | 24 | 429.6 | 55.9 | 66.9 (3.2–91.7) |
| Asthma | 485 | 9 | 236.8 | 38.0 | 929 | 27 | 454.4 | 59.4 | 37.5 (-37.2–74.2) |
| Chronic respiratory/pulmonary disease | 889 | 13 | 440.5 | 29.5 | 1720 | 49 | 846.7 | 57.9 | 50.0 (6.2–75.2) |
| Chronic heart failure | 162 | 1 | 81.9 | 12.2 | 312 | 6 | 158.9 | 37.7 | 72.1 (-131.9–99.4) |
| Diabetes mellitus type 1 or 2 | 1141 | 10 | 568.8 | 17.6 | 2295 | 34 | 1142.0 | 29.8 | 41.4 (-21.4–74.2) |
| Advanced liver or renal disease | 284 | 5 | 145.8 | 34.3 | 560 | 7 | 289.1 | 24.2 | -34.5 (-394.4–66.5) |

Results based on post-hoc exploratory analyses, except for those in the overall trial population. Analysis included cases collected from day 15 post-dose 2 until end of Northern Hemisphere season 2 (efficacy data lock point: 31 March 2023) from participants who received adjuvanted RSVPreF3 as dose 1 and placebo as dose 2 and from participants who received placebo as doses 1 and 2. Vaccine efficacy was estimated using a Poisson model adjusted for age and region.

Adjuvanted RSVPreF3, adjuvanted respiratory syncytial virus (RSV) prefusion F protein-based vaccine; RSV-LRTD, RSV-related lower respiratory tract disease confirmed by the adjudication committee; RSV-ARI, RSV-related acute respiratory illness; N, number of participants in the dose 2 modified exposed population; n, number of participants with ≥1 RSV-LRTD or RSV-ARI; T, sum of follow-up time from day 15 post-dose 2 until first occurrence of the event, data lock point, or drop-out; IR, incidence rate of participants reporting at least one event, expressed per 1000 person-years (py), calculated as 1000 x (n/T); CI, confidence interval; COPD, chronic obstructive pulmonary disease; BMI, body mass index (18.5–<25 kg/m$^2$: healthy weight; 25–<30 kg/m$^2$: overweight; ≥30 kg/m$^2$: obesity).

[a]More than one type of pre-existing condition of interest could occur in a single participant. [b]Among the 50 (adjuvanted RSVPreF3) and 112 (placebo) participants with a BMI <18.5 kg/m$^2$, none reported an RSV-LRTD.

**Supplementary Table 4. Vaccine efficacy of a single adjuvanted RSVPreF3 dose against first occurrence of RSV-LRTD and RSV-ARI during the third RSV season, overall and among participants with pre-existing medical conditions (dose 2 modified exposed population)**

| Endpoint | Adjuvanted RSVPreF3 group (single dose) | | | | Placebo group | | | | Vaccine efficacy, % |
|---|---|---|---|---|---|---|---|---|---|
| Subgroup | N | n | T, py | IR per 1000 py | N | n | T, py | IR per 1000 py | (95% CI) |
| **RSV-LRTD** | | | | | | | | | |
| Overall | 4988 | 16 | 2717.2 | 5.9 | 10,031 | 61 | 5407.4 | 11.3 | 48.0 (8.7–72.0) |
| By pre-existing condition of interest[a] | | | | | | | | | |
| COPD | 464 | 3 | 245.4 | 12.2 | 881 | 10 | 457.9 | 21.8 | 40.1 (-134.6–89.5) |
| Asthma | 494 | 2 | 270.2 | 7.4 | 946 | 14 | 508.9 | 27.5 | 72.1 (-21.6–96.9) |
| Chronic respiratory/pulmonary disease | 903 | 5 | 484.3 | 10.3 | 1744 | 23 | 921.6 | 25.0 | 58.1 (-12.8–87.6) |
| Chronic heart failure | 166 | 0 | 81.7 | 0.0 | 317 | 1 | 167.1 | 6.0 | 100 (-3879.3–100) |
| Diabetes mellitus type 1 or 2 | 1144 | 2 | 613.1 | 3.3 | 2304 | 16 | 1227.5 | 13.0 | 74.8 (-7.3–97.2) |
| Advanced liver or renal disease | 289 | 2 | 149.9 | 13.3 | 573 | 4 | 305.1 | 13.1 | -2.4 (-620.4–90.8) |
| By BMI category[b] | | | | | | | | | |
| BMI 18.5–<25 kg/m$^2$ | 1192 | 2 | 650.8 | 3.1 | 2402 | 8 | 1294.8 | 6.2 | 50.7 (-147.5–94.9) |
| BMI 25–<30 kg/m$^2$ | 1842 | 6 | 1012.3 | 5.9 | 3705 | 21 | 2008.7 | 10.5 | 43.3 (-45.3–81.3) |
| BMI ≥30 kg/m$^2$ | 1899 | 8 | 1024.9 | 7.8 | 3809 | 32 | 2044.9 | 15.6 | 50.4 (-10.0–80.3) |

| | | | | | | | | | |
|---|---|---|---|---|---|---|---|---|---|
| **RSV-ARI** | | | | | | | | | |
| Overall | 4988 | 30 | 2712.6 | 11.1 | 10,031 | 113 | 5391.5 | 21.0 | 47.4 (20.7–66.1) |
| By pre-existing condition of interest[a] | | | | | | | | | |
| COPD | 464 | 3 | 245.4 | 12.2 | 881 | 13 | 457.1 | 28.4 | 55.1 (-64.6– 91.8) |
| Asthma | 494 | 5 | 269.1 | 18.6 | 946 | 18 | 507.7 | 35.5 | 45.1 (-53.8–84.1) |
| Chronic respiratory/pulmonary disease | 903 | 8 | 483.2 | 16.6 | 1744 | 29 | 919.9 | 31.5 | 47.2 (-18.6–79.2) |
| Chronic heart failure | 166 | 1 | 81.4 | 12.3 | 317 | 2 | 166.8 | 12.0 | -11.1 (-2042.8–98.1) |
| Diabetes mellitus type 1 or 2 | 1144 | 4 | 612.7 | 6.5 | 2304 | 24 | 1224.8 | 19.6 | 66.7 (3.1–91.6) |
| Advanced liver or renal disease | 289 | 2 | 149.9 | 13.3 | 573 | 8 | 303.9 | 26.3 | 50.3 (-150.5–94.9) |
| By BMI category[b] | | | | | | | | | |
| BMI 18.5–<25 kg/$m^2$ | 1192 | 8 | 648.8 | 12.3 | 2402 | 23 | 1290.1 | 17.8 | 30.4 (-61.3–73.1) |
| BMI 25–<30 kg/$m^2$ | 1842 | 11 | 1010.5 | 10.9 | 3705 | 43 | 2001.7 | 21.5 | 50.0 (1.3–76.7) |
| BMI ≥30 kg/$m^2$ | 1899 | 11 | 1024.1 | 10.7 | 3809 | 45 | 2041.1 | 22.0 | 50.8 (3.3–77.1) |

Results based on post-hoc exploratory analyses, except for those in the overall trial population. Analysis included cases collected from the start of Northern Hemisphere season 3 (1 October 2023) until the end of Northern Hemisphere season 3 (efficacy data lock point: 30 April 2024) from participants who received adjuvanted RSVPreF3 as dose 1 and placebo as dose 2 and from participants who received placebo as doses 1 and 2. Vaccine efficacy was estimated using a Poisson model adjusted for age and region.

Adjuvanted RSVPreF3, adjuvanted respiratory syncytial virus (RSV) prefusion F protein-based vaccine; RSV-LRTD, RSV-related lower respiratory tract disease confirmed by the adjudication committee; RSV-ARI, RSV-related acute respiratory illness; N, number of participants in the dose 2 modified exposed population; n, number of participants with ≥1 RSV-LRTD or RSV-ARI; T, sum of follow-up time from start of season 3 until first occurrence of the event, data lock point, or drop-out; IR, incidence rate of participants reporting at least one event, expressed per 1000 person-years (py), calculated as 1000 x (n/T); CI, confidence interval; COPD, chronic obstructive pulmonary disease; BMI, body mass index (18.5–<25 kg/$m^2$: healthy weight; 25–<30 kg/$m^2$: overweight; ≥30 kg/$m^2$: obesity).

[a]More than one type of pre-existing condition of interest could occur in a single participant. [b]Among the 50 (adjuvanted RSVPreF3) and 112 (placebo) participants with a BMI <18.5 kg/$m^2$, none reported an RSV-LRTD and two (placebo) reported an RSV-ARI.

**Supplementary Table 5. Vaccine efficacy of a first adjuvanted RSVPreF3 dose followed by a second dose 1 year later (revaccination) against first occurrence of RSV-LRTD and RSV-ARI over three RSV seasons, overall and among participants with pre-existing medical conditions (modified exposed population)**

| Endpoint | Adjuvanted RSVPreF3 (revaccination) | | | | Placebo | | | | Vaccine efficacy, % |
|---|---|---|---|---|---|---|---|---|---|
| Subgroup | N | n | T, py | IR per 1000 py | N | n | T, py | IR per 1000 py | (CI[a]) |
| **RSV-LRTD** | | | | | | | | | |
| Overall | 12,468 | 38 | 18,744.4 | 2.0 | 12,498 | 192 | 25,468.3 | 7.5 | 67.8 (51.8–79.1) |
| By pre-existing condition of interest[b] | | | | | | | | | |
| No condition | 7454 | 20 | 11,283.8 | 1.8 | 7547 | 91 | 15,558.9 | 5.8 | 64.2 (41.1–79.2) |
| ≥1 condition | 5014 | 18 | 7460.6 | 2.4 | 4951 | 101 | 9909.4 | 10.2 | 71.2 (51.9–83.7) |
| ≥1 cardiorespiratory condition | 2577 | 11 | 3793.9 | 2.9 | 2504 | 73 | 4964.0 | 14.7 | 75.9 (54.1–88.5) |
| COPD | 1181 | 3 | 1720.6 | 1.7 | 1161 | 35 | 2265.0 | 15.5 | 86.1 (55.5–97.3) |
| Asthma | 1226 | 9 | 1836.6 | 4.9 | 1160 | 41 | 2372.5 | 17.3 | 63.8 (23.8–84.7) |
| Chronic respiratory/pulmonary disease | 2287 | 11 | 3391.9 | 3.2 | 2200 | 72 | 4405.8 | 16.3 | 75.7 (53.7–88.5) |
| Chronic heart failure | 427 | 0 | 585.4 | 0.0 | 425 | 4 | 782.4 | 5.1 | 100.0 (-53.7–100.0) |
| ≥1 endocrine/metabolic condition | 3243 | 9 | 4846.6 | 1.9 | 3274 | 49 | 6590.0 | 7.4 | 69.9 (37.5–87.1) |
| Diabetes mellitus type 1 or 2 | 2856 | 8 | 4276.4 | 1.9 | 2898 | 44 | 5831.9 | 7.5 | 70.5 (36.2–88.1) |

| | | | | | | | | | |
|---|---|---|---|---|---|---|---|---|---|
| Advanced liver or renal disease | 698 | 1 | 1021.6 | 1.0 | 710 | 10 | 1426.4 | 7.0 | 82.1 (-29.2–99.6) |
| By BMI category[c] | | | | | | | | | |
| BMI 18.5–<25 kg/m$^2$ | 3051 | 7 | 4602.4 | 1.5 | 2999 | 32 | 6127.3 | 5.2 | 63.9 (16.1–86.6) |
| BMI 25–<30 kg/m$^2$ | 4560 | 21 | 6873.7 | 3.1 | 4600 | 63 | 9356.9 | 6.7 | 45.1 (8.2–68.3) |
| BMI ≥30 kg/m$^2$ | 4716 | 10 | 7071.9 | 1.4 | 4745 | 97 | 9689.9 | 10.0 | 83.2 (67.6–92.2) |
| **RSV-ARI** | | | | | | | | | |
| Overall | 12,468 | 103 | 18.678.5 | 5.5 | 12,498 | 385 | 25,236.9 | 15.3 | 58.4 (48.1–67.0) |
| By pre-existing condition of interest[b] | | | | | | | | | |
| No condition | 7454 | 63 | 11,238.5 | 5.6 | 7547 | 212 | 15,411.4 | 13.8 | 53.4 (37.9–65.5) |
| ≥1 condition | 5014 | 40 | 7440.0 | 5.4 | 4951 | 173 | 9825.5 | 17.6 | 64.6 (49.7–75.7) |
| ≥1 cardiorespiratory condition | 2577 | 21 | 3785.3 | 5.5 | 2504 | 106 | 4924.5 | 21.5 | 69.7 (51.0–82.1) |
| COPD | 1181 | 5 | 1719.4 | 2.9 | 1161 | 50 | 2242.9 | 22.3 | 85.1 (62.6–95.4) |
| Asthma | 1226 | 16 | 1830.0 | 8.7 | 1160 | 53 | 2357.3 | 22.5 | 51.3 (12.8–74.2) |
| Chronic respiratory/pulmonary disease | 2287 | 20 | 3384.1 | 5.9 | 2200 | 98 | 4372.4 | 22.4 | 68.5 (48.3–81.6) |
| Chronic heart failure | 427 | 1 | 584.6 | 1.7 | 425 | 11 | 776.2 | 14.2 | 87.1 (8.0–99.7) |
| ≥1 endocrine/metabolic condition | 3243 | 23 | 4833.5 | 4.8 | 3274 | 100 | 6530.9 | 15.3 | 65.0 (44.3–78.9) |
| Diabetes mellitus type 1 or 2 | 2856 | 21 | 4263.7 | 4.9 | 2898 | 89 | 5776.5 | 15.4 | 64.5 (42.0–79.1) |
| Advanced liver or renal disease | 698 | 3 | 1021.1 | 2.9 | 710 | 25 | 1412.9 | 17.7 | 80.3 (34.5–96.2) |

| | | | | | | | | | |
|---|---|---|---|---|---|---|---|---|---|
| By BMI category[c] | | | | | | | | | |
| BMI 18.5–<25 kg/m$^2$ | 3051 | 29 | 4581.7 | 6.3 | 2999 | 83 | 6066.1 | 13.7 | 46.1 (16.5–66.1) |
| BMI 25–<30 kg/m$^2$ | 4560 | 42 | 6850.1 | 6.1 | 4600 | 133 | 9278.4 | 14.3 | 49.9 (28.3–65.6) |
| BMI ≥30 kg/m$^2$ | 4716 | 30 | 7050.7 | 4.3 | 4745 | 167 | 9598.5 | 17.4 | 72.5 (59.1–82.1) |

Results based on post-hoc exploratory analyses, except for those in the overall trial population, in participants with no condition, ≥1 condition, ≥1 cardiorespiratory, and ≥1 endocrine/metabolic condition of interest. Analysis included data collected from day 15 post-dose 1 until end of Northern Hemisphere season 3 (efficacy data lock point: 30 April 2024) in participants who did not receive dose 3, or until dose 3 administration, or until end of Southern Hemisphere season 1 (efficacy data lock point: 30 September 2022) for participants who did not receive dose 2. Participants who received adjuvanted RSVPreF3 as doses 1 and 2 (RSV revaccination group) and participants in the placebo group contributed to the entire follow-up. Participants who received adjuvanted RSVPreF3 as dose 1 and placebo as dose 2 (RSV single-dose group) contributed to season 1 but were censored at dose 2. Vaccine efficacy was estimated using a Poisson model adjusted for age, region, and season.

Adjuvanted RSVPreF3, adjuvanted respiratory syncytial virus (RSV) prefusion F protein-based vaccine; RSV-LRTD, RSV-related lower respiratory tract disease confirmed by the adjudication committee; RSV-ARI, RSV-related acute respiratory illness; N, number of participants in the modified exposed population; n, number of participants with ≥1 RSV-LRTD or RSV-ARI; T, sum of follow-up time (from day 15 post-dose 1 until first occurrence of the event, data lock point, drop-out, or dose 3 administration); IR, incidence rate of participants reporting at least one event, expressed per 1000 person-years (py), calculated as 1000 x (n/T); CI, confidence interval; COPD, chronic obstructive pulmonary disease; BMI, body mass index (18.5–<25 kg/m$^2$: healthy weight; 25–<30 kg/m$^2$: overweight; ≥30 kg/m$^2$: obesity).

[a]97.5% CI for RSV-LRTD, overall (confirmatory secondary endpoint); 95% CI for other endpoints. [b]More than one type of pre-existing condition of interest could occur in a single participant. [c]Among the 131 (adjuvanted RSVPreF3) and 145 (placebo) participants with a BMI <18.5 $kg/m^2$, none reported an RSV-LRTD, and two (adjuvanted RSVPreF3) and two (placebo) reported an RSV-ARI.

**Supplementary Table 6. Occurrence of RSV-ARI-related complications over three RSV seasons (modified exposed population)**

| Complication | RSV single-dose group (N=6225) | Placebo group (N=12,498) |
|---|---|---|
| At least one complication | 11 (0.2) | 50 (0.4) |
| At least one respiratory complication | 11 (0.2) | 48 (0.4) |
| Pneumonia | 3 (0.0) | 8 (0.1) |
| New diagnosis of COPD | 0 (0.0) | 1 (0.0) |
| Exacerbation of COPD | 3 (0.0) | 13 (0.1) |
| New diagnosis of asthma | 0 (0.0) | 0 (0.0) |
| Exacerbation of asthma | 2 (0.0) | 12 (0.1) |
| Other respiratory complication[a] | 3 (0.0) | 16 (0.1) |
| At least one non-respiratory complication | 0 (0.0) | 3 (0.0) |
| New-onset congestive heart failure | 0 (0.0) | 1 (0.0) |
| Worsening congestive heart failure | 0 (0.0) | 0 (0.0) |
| Myocardial infarction | 0 (0.0) | 0 (0.0) |
| Stroke | 0 (0.0) | 0 (0.0) |
| Diabetes | 0 (0.0) | 0 (0.0) |
| Other non-respiratory complication[b] | 0 (0.0) | 2 (0.0) |

Results based on post-hoc exploratory analyses. Data are n (%), with n being the number of participants reporting at least one of the indicated complications and % calculated as n/N. Analysis included data collected from day 15 post-dose 1 until end of Northern Hemisphere season 3 (efficacy data lock point: 30 April 2024) or until end of Southern Hemisphere season 1 (efficacy data lock point: 30 September 2022) for participants who did not receive dose 2.

RSV-ARI, respiratory syncytial virus (RSV)-related acute respiratory illness; N, number of participants per group in the modified exposed population; COPD, chronic obstructive pulmonary disease.

[a]Other respiratory complications reported were acute bronchitis, bacterial bronchitis, bronchitis, exacerbation of bronchiectasis, exacerbation of reactive airway disease, RSV infection, sinusitis, viral lower respiratory tract infection, worsening of dyspnoea, worsening of viral bronchitis, whooping cough, and chest infection. [b]Other non-respiratory complications reported were acute otitis media.

**Supplementary Table 7. Vaccine efficacy of a single adjuvanted RSVPreF3 dose against RSV-ARI-related complications and disease exacerbations during the first RSV season and over the first two seasons, overall and among participants with pre-existing medical conditions (modified exposed population)**

| Endpoint | Adjuvanted RSVPreF3 group (single dose) | | | | Placebo group | | | | Vaccine efficacy, % |
|---|---|---|---|---|---|---|---|---|---|
| Subgroup | N | n | T, py | IR per 1000 py | N | n | T, py | IR per 1000 py | (95% CI) |
| **RSV season 1** | | | | | | | | | |
| **RSV-ARI-related complications** | | | | | | | | | |
| Overall | 12,468 | 4 | 11,695.7 | 0.3 | 12,498 | 16 | 11,680.4 | 1.4 | 75.1 (22.7−93.9) |
| By pre-existing condition of interest[a] | | | | | | | | | |
| No condition | 7454 | 1 | 6939.8 | 0.1 | 7547 | 5 | 7022.4 | 0.7 | 79.4 (-84.0−99.6) |
| ≥1 condition | 5014 | 3 | 4755.8 | 0.6 | 4951 | 11 | 4658.0 | 2.4 | 73.5 (-0.2−95.3) |
| ≥1 cardiorespiratory condition | 2577 | 2 | 2449.3 | 0.8 | 2504 | 11 | 2357.8 | 4.7 | 82.5 (20.0−98.1) |
| COPD | 1181 | 0 | 1125.9 | 0.0 | 1161 | 7 | 1086.3 | 6.4 | 100 (47.3−100) |
| Asthma | 1226 | 2 | 1175.2 | 1.7 | 1160 | 4 | 1109.7 | 3.6 | 53.0 (-228.3−95.8) |
| Chronic respiratory/pulmonary disease | 2287 | 2 | 2182.3 | 0.9 | 2200 | 11 | 2081.3 | 5.3 | 82.7 (20.7−98.1) |
| Chronic heart failure | 427 | 0 | 396.1 | 0.0 | 425 | 2 | 389.4 | 5.1 | 100 (-246.3−100) |

| | | | | | | | | | |
|---|---|---|---|---|---|---|---|---|---|
| **RSV-ARI-related exacerbation of COPD** | | | | | | | | | |
| By pre-existing condition of interest | | | | | | | | | |
| COPD | 1181 | 0 | 1125.9 | 0.0 | 1161 | 6 | 1087.3 | 5.5 | 100 (37.4–100) |
| **RSV-ARI-related exacerbation of asthma** | | | | | | | | | |
| By pre-existing condition of interest | | | | | | | | | |
| Asthma | 1226 | 2 | 1175.2 | 1.7 | 1160 | 3 | 1110.5 | 2.7 | 37.9 (-442.9–94.8) |
| **RSV seasons 1+ 2** | | | | | | | | | |
| **RSV-ARI-related complications** | | | | | | | | | |
| Overall | 12,468 | 10 | 16,896.4 | 0.6 | 12,498 | 38 | 21,825.6 | 1.7 | 61.9 (21.4–83.2) |
| By pre-existing condition of interest[a] | | | | | | | | | |
| No condition | 7454 | 2 | 10,093.8 | 0.2 | 7547 | 9 | 13,229.8 | 0.7 | 69.9 (-48.1–96.9) |
| ≥1 condition | 5014 | 8 | 6802.7 | 1.2 | 4951 | 29 | 8595.8 | 3.4 | 60.1 (9.6–84.4) |
| ≥1 cardiorespiratory condition | 2577 | 7 | 3479.7 | 2.0 | 2504 | 27 | 4328.8 | 6.2 | 63.4 (12.9–86.7) |
| COPD | 1181 | 2 | 1597.9 | 1.3 | 1161 | 15 | 1968.0 | 7.6 | 79.9 (11.4–97.8) |
| Asthma | 1226 | 5 | 1685.8 | 3.0 | 1160 | 14 | 2055.9 | 6.8 | 50.3 (-48.0–86.2) |
| Chronic respiratory/pulmonary disease | 2287 | 7 | 3106.7 | 2.3 | 2200 | 27 | 3825.7 | 7.1 | 63.6 (13.3–86.8) |
| Chronic heart failure | 427 | 0 | 555.9 | 0.0 | 425 | 2 | 700.6 | 2.9 | 100 (-245.4–100) |

| | | | | | | | | | |
|---|---|---|---|---|---|---|---|---|---|
| **RSV-ARI-related exacerbation of COPD** | | | | | | | | | |
| By pre-existing condition of interest | | | | | | | | | |
| COPD | 1181 | 2 | 1597.9 | 1.3 | 1161 | 11 | 1972.2 | 5.6 | 73.9 (-23.7−97.2) |
| **RSV-ARI-related exacerbation of asthma** | | | | | | | | | |
| By pre-existing condition of interest | | | | | | | | | |
| Asthma | 1226 | 3 | 1687.2 | 1.8 | 1160 | 9 | 2060.9 | 4.4 | 57.9 (-71.6−92.8) |

Results based on post-hoc exploratory analyses, except for those in the overall trial population. Complications could include respiratory complications (pneumonia, new diagnosis or exacerbation of COPD or asthma, other respiratory complications [e.g., sinusitis, bronchitis, dyspnoea]) and non-respiratory complications (new onset or worsening of congestive heart failure, myocardial infarction, stroke, diabetes, other non-respiratory complications). Analysis for season 1 included data collected from day 15 post-dose 1 until end of Southern Hemisphere season 1 (efficacy data lock point: 30 September 2022) or up to dose 2 administration. Analysis over the first two seasons included data collected from day 15 post-dose 1 until end of Southern Hemisphere season 2 (efficacy data lock point: 30 September 2023) or up to dose 3 administration, or up to end of Southern Hemisphere season 1 (efficacy data lock point: 30 September 2022) if no dose 2 administration; participants who received adjuvanted RSVPreF3 as dose 1 and placebo as dose 2 (RSV single-dose group) and participants in the placebo group contributed to the entire follow-up; participants who received adjuvanted RSVPreF3 as doses 1 and 2 (RSV revaccination group) contributed to season 1 but were censored at dose 2. Vaccine efficacy was estimated using a Poisson model adjusted for age and region (for season 1 analysis), or age, region, and season (for analysis over two seasons).

Adjuvanted RSVPreF3, adjuvanted respiratory syncytial virus (RSV) prefusion F protein-based vaccine; RSV-ARI, RSV-related acute respiratory illness; N, number of participants in the modified exposed population; n, number of participants with ≥1 RSV-ARI-related complication; T, sum of follow-up time (from day 15 post-dose 1 until first occurrence of the event, data lock point, drop-out, or dose 2 administration [for season 1 analysis], or dose 3 administration [for analysis over two seasons]); IR, incidence rate of participants reporting at least one event, expressed per 1000 person-years (py), calculated as 1000 x (n/T); CI, confidence interval; COPD, chronic obstructive pulmonary disease.

[a]More than one type of pre-existing condition of interest could occur in a single participant.

**Supplementary Figure 1. RSV-A and RSV-B neutralising titres pre-dose 1, one month post-dose 1, pre-season 2, and pre-season 3, by pre-existing medical conditions of interest (per-protocol population)**

**A. RSV-A neutralising titres**

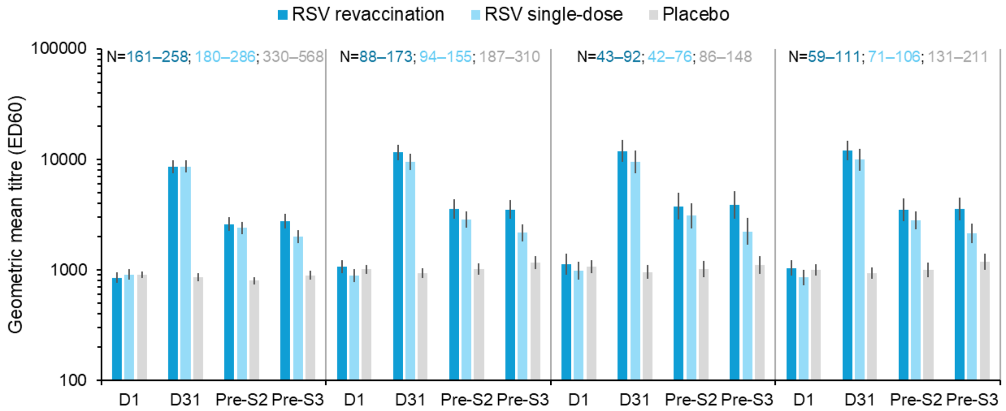


| MGI vs D1 (95% CI) | | No condition | ≥1 condition | ≥1 cardiorespiratory condition | ≥1 endocrine/metabolic condition |
|---|---|---|---|---|---|
| RSV revaccination | D31 | 10.0 (8.9–11.4) | 11.0 (9.1–13.3) | 10.9 (8.4–14.2) | 11.7 (9.0–15.2) |
| | Pre-S2 | 3.0 (2.7–3.4) | 3.4 (2.8–4.0) | 3.4 (2.7–4.4) | 3.4 (2.7–4.3) |
| | Pre-S3 | 3.4 (3.1–3.8) | 3.5 (2.8–4.2) | 3.5 (2.6–4.8) | 3.5 (2.8–4.5) |
| RSV single-dose | D31 | 9.4 (8.3–10.7) | 10.8 (9.0–13.1) | 9.9 (7.7–12.7) | 11.6 (9.2–14.8) |
| | Pre-S2 | 2.6 (2.4–2.9) | 3.1 (2.7–3.6) | 3.1 (2.6–3.8) | 3.2 (2.6–3.8) |
| | Pre-S3 | 2.3 (2.1–2.6) | 2.4 (2.0–2.8) | 2.4 (2.0–2.9) | 2.2 (1.8–2.7) |
| Placebo | D31 | 1.0 (0.9–1.0) | 0.9 (0.9–1.0) | 0.9 (0.8–1.0) | 0.9 (0.9–1.0) |
| | Pre-S2 | 0.9 (0.9–1.0) | 1.0 (0.9–1.1) | 1.0 (0.9–1.1) | 1.0 (0.9–1.1) |
| | Pre-S3 | 1.0 (1.0–1.1) | 1.1 (1.0–1.3) | 1.1 (1.0–1.3) | 1.2 (1.0–1.3) |

**B. RSV-B neutralising titres**

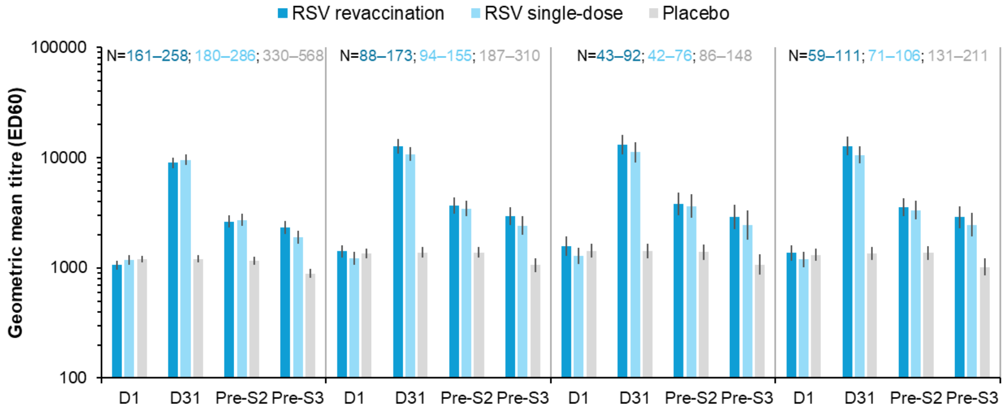


| MGI vs D1 (95% CI) | | No condition | ≥1 condition | ≥1 cardiorespiratory condition | ≥1 endocrine/metabolic condition |
|---|---|---|---|---|---|
| RSV revaccination | D31 | 8.5 (7.5–9.5) | 9.1 (7.7–10.7) | 8.4 (6.6–10.7) | 9.3 (7.5–11.7) |
| | Pre-S2 | 2.6 (2.3–2.9) | 2.6 (2.2–3.1) | 2.4 (1.9–3.1) | 2.6 (2.1–3.3) |
| | Pre-S3 | 2.4 (2.1–2.7) | 2.4 (2.0–2.8) | 2.1 (1.6–2.8) | 2.4 (1.9–3.0) |
| RSV single-dose | D31 | 8.1 (7.2–9.1) | 8.9 (7.7–10.4) | 8.8 (7.2–10.7) | 9.0 (7.4–11.0) |
| | Pre-S2 | 2.3 (2.1–2.6) | 2.7 (2.4–3.1) | 2.7 (2.2–3.2) | 2.7 (2.3–3.2) |
| | Pre-S3 | 1.6 (1.5–1.8) | 1.9 (1.6–2.1) | 1.7 (1.5–2.1) | 1.9 (1.6–2.3) |
| Placebo | D31 | 1.0 (1.0–1.1) | 1.0 (1.0–1.1) | 1.0 (1.0–1.1) | 1.0 (1.0–1.1) |
| | Pre-S2 | 1.0 (0.9–1.0) | 1.1 (1.0–1.1) | 1.0 (0.9–1.1) | 1.1 (1.0–1.2) |
| | Pre-S3 | 0.8 (0.7–0.8) | 0.8 (0.7–0.9) | 0.8 (0.7–0.9) | 0.8 (0.7–0.9) |

Error bars depict 95% CIs. The lower limits of quantification were 18 ED60 for the RSV-A neutralisation assay and 30 ED60 for the RSV-B neutralisation assay.

RSV, respiratory syncytial virus; RSV revaccination, group of participants who received a first dose of adjuvanted RSV prefusion F protein-based vaccine (adjuvanted RSVPreF3) pre-season 1 and a second dose (revaccination) pre-season 2; RSV single-dose, group of participants who received a single dose of adjuvanted RSVPreF3 pre-season 1 and a placebo dose pre-season 2; placebo, group of participants who received placebo pre-season 1 and pre-season 2; N, number of participants with available results (range across timepoints is given per group and per underlying condition subgroup); ED60, estimated dilution 60; D1, day 1 (pre-dose 1); D31, day 31 (1 month post-dose 1); pre-S2, pre-RSV season 2 (pre-dose 2); pre-S3, pre-RSV season 3; MGI, mean geometric increase (i.e., geometric mean of the within-participant ratios of the post-vaccination over pre-vaccination titres); CI, confidence interval.
